# Genome-wide association study in Brazil identifies genetic susceptibility to tuberculosis with single-cell gene effects

**DOI:** 10.1101/2025.03.13.25323932

**Authors:** Kimberly A Dill-McFarland, Bruno B Andrade, Marina C Figueiredo, Alice M S Andrade, Francys Avendaño-Rangel, Marcelo Cordeiro-Santos, Afrânio L Kritski, Valeria C Rolla, Juan M Cubillos-Angulo, Spyros A Kalams, Josh D Simmons, Jared M Oakes, Jonathan Peña Avila, Helder I Nakaya, Rama D Gangula, Peter F Rebeiro, Gustavo Amorim, Simon A Mallal, Regional Prospective Observational Research in Tuberculosis (RePORT)-Brazil Consortium, Timothy R Sterling, Thomas R Hawn

**Affiliations:** Allergy and Infectious Diseases, Department of Medicine, University of Washington, Seattle, WA, USA; Laboratório de Pesquisa Clínica e Translacional, Instituto Gonçalo Moniz, Fundação Oswaldo Cruz (FIOCRUZ), Salvador, BA, Brazil; Division of Infectious Diseases, School of Medicine, Johns Hopkins University, Baltimore, MD, USA; Division of Infectious Diseases, Department of Medicine, Vanderbilt University Medical Center, Nashville, TN, USA; Fundação de Medicina Tropical Dr. Heitor Vieira Dourado do Amazonas, Manaus, AM, Brazil; Curso de Medicina, Universidade do Estado do Amazonas (UEA), Manaus, AM, Brazil; Programa Acadêmico de Tuberculose da Faculdade de Medicina, Universidade Federal do Rio de Janeiro, Rio de Janeiro, RJ, Brazil; Laboratório de Pesquisa Clínica em Micobacteriose, Instituto Nacional de Infectologia Evandro Chagas (FIOCRUZ), Rio de Janeiro, RJ, Brazil; Fundación Universitaria de Ciencias de la Salud, Bogotá D.C., Colombia; Department of Pathology, Microbiology, and Immunology, Vanderbilt University Medical Center, Nashville, TN, USA; Department of Clinical and Toxicological Analyses, University of São Paulo, São Paulo, SP, Brazil; Hospital Israelita Albert Einstein, São Paulo, SP, Brazil; Department of Biostatistics, Vanderbilt University Medical Center, Nashville, TN, USA

## Abstract

**Background:** Although genetic factors contribute to tuberculosis (TB) risk, no cross-population causal variants have been identified by genome-wide association studies (GWAS).

**Methods:** Here, we utilized low-pass whole genome sequencing (lpWGS) with high-pass WGS verified imputation plus detailed epidemiologic risk factors and single-cell expression quantitative loci (sceQTL) to address prior GWAS limitations.

**Results:** Using 947 pulmonary tuberculosis (PTB) cases and 1807 close contact controls in the Regional Prospective Observational Research in TB (RePORT) study in Brazil, we estimated PTB heritability to be 40 - 68%. We identified 17 SNPs associated with PTB (P<5E-8) after adjustment for major risk factors (HIV, diabetes, smoking). Seven of these SNPs were associated with peripheral blood cell-specific sceQTLs in controls. Specifically, SNPs cis to transcription factors ZNF717 and MAML3 were associated with PTB disease and gene expression in myeloid, T cells, or B cells. The nucleoporin-associated genes NUP93 and AGFG1 were also associated with sceQTLs in multiple cell types including dendritic, natural killer, or T cells.

**Conclusion:** Overall, this study utilized lpWGS, in-depth epidemiology, and single-cell analyses to detect population-specific genetic risk factors for PTB in Brazil.

**SUMMARY:** Robust correction for tuberculosis risk factors in GWAS in combination with paired single-cell transcriptomics reveals novel genetic risk of pulmonary tuberculosis with measurable consequences for baseline gene expression in multiple cell types.

## INTRODUCTION

Multiple studies over the past 50 years suggest that host genetics regulate susceptibility to tuberculosis (TB), yet the major causal genes remain largely unknown (1, 2). This gap in knowledge regarding “missing heritability” is exemplified by the lack of reproducible findings among genome-wide association studies (GWAS) of TB in several countries (3–12). The reasons for missing heritability in TB GWAS are multifactorial. Differences in underlying population genetics, population-specific genetic risk factors, genotyping platforms, imputation reference panels, Mycobacterium tuberculosis (Mtb) strains, and a lack of adjustment for clinical TB risk factors may hinder the reproducibility of TB GWAS results. In addition, prior GWAS often lack direct experimental connections of associated genetic variants to functional outcomes that may impact TB outcomes. Designing studies to address these limitations may foster the discovery of the missing genetic regulators of susceptibility to TB.

Genotyping methods for GWAS studies continue to evolve to better measure high-confidence genetic variants within and across populations. High-pass whole genome sequencing (hpWGS) with at least 30X coverage is the gold-standard to comprehensively define genetic variation that may be population- and/or disease-specific. However, due to budgetary constraints, alternative methods are more often employed. Single nucleotide polymorphism (SNP) arrays with or without imputation are the most common genotyping platforms. These platforms were used in all prior TB GWAS, but they rely on pre-selected SNPs based on previously genetically characterized populations. Low-pass WGS (lpWGS, ≤ 1X coverage), along with recent advances in imputation, offers a cost-effective alternative to capture more population- and disease-specific variation compared to SNP arrays (13). In addition, while the discovery of statistically significant SNPs remains the primary goal of GWAS, evidence of the functional effects of these SNPs is needed to fully elucidate their role in TB pathogenesis. In some studies, database-derived expression quantitative trail loci (eQTL) provide evidence of SNP function, but current transcriptomic databases are limited by the small number of cell types and/or human populations available for comparison to GWAS studies. Single-cell eQTL data in the same population as a GWAS could address this gap and expand the functional assessment of candidate GWAS SNPs.

In addition, precise phenotyping of traits is an essential feature of genetic studies. For TB cases, genetic signals may be obscured by other strong TB risk factors including HIV, diabetes mellitus (DM), smoking, alcohol consumption, malnutrition, and others. In addition, control misclassification can occur if there is insufficient Mtb exposure and/or follow-up duration among controls. Previous TB GWAS mainly focused on pulmonary TB (PTB) in HIV-negative populations with adjustments for age, sex, and ancestry but not other risk factors such as DM and smoking (3–12). The method to control for potential confounders is most often a matched case-control design while an alternative full cohort, covariate-adjusted method is underexplored despite providing more statistical power. Full cohort GWAS may be particularly powerful in highly admixed populations where higher genetic diversity and more complex linkage disequilibrium structures necessitate larger sample sizes to achieve robust statistical findings (14). Brazil represents one such highly admixed population with high TB rates (49 cases per 100,000 population (15)), nearly 100% bacillus Calmette-Guérin (BCG) vaccination (16), and significant variability in disease burden by region (16). The causes of regional differences remain largely unknown but most commonly are attributed to access to healthcare, socioeconomic indicators, or climate (16), leaving underlying population genetics unexplored. Despite practices since 2018 to administer TB infection preventative care to all contacts regardless of HIV or tuberculin skin test (TST) status as well as incorporate interferon-gamma release assay (IGRA) results in diagnosis (17), TB rates have remained relatively stable across Brazil (18).

To address some of the methodologic limitations of prior TB GWAS, we prospectively enrolled PTB cases and their close contacts in the Regional Prospective Observational Research in TB (RePORT) study in Brazil, genotyped SNPs by lpWGS with imputation, validated imputation by 30X hpWGS, and performed both full cohort covariate-adjusted and risk-controlled sub-cohort analyses. We also assessed the functional relevance of candidate SNPs through eQTL analysis in a peripheral blood mononuclear cell (PBMC) single-cell RNA-seq substudy in RePORT-Brazil. Together, GWAS and eQTL analyses were used to determine the genetic determinants of active PTB.

## RESULTS

### GWAS study design

To discover genetic determinants of pulmonary tuberculosis (PTB) susceptibility in Brazil, we examined genotypes by low-pass whole genome sequencing (lpWGS) with imputation and high-pass WGS (hpWGS) confirmation in culture-confirmed PTB cases and close contact controls from the RePORT-Brazil cohort (Figure 1A, Supplemental Figure 1). We then performed two complementary analyses: a full cohort risk-adjusted design (N = 947 cases, 1807 controls) to maximize statistical power to detect smaller effects and a risk-controlled sub-cohort strategy (N = 578 cases, 241 controls) to reduce PTB risk factor variance in order to detect less biased causal effects. When compared to controls in the full cohort (Table 1), PTB cases had a higher male frequency (67% cases vs 41% controls, P = 1E-37), were older (38 ± 14 vs 33 ± 19, P = 2E-14), had lower BMI (21 ± 3.5 vs 25 ± 6.5, P = 7E-71), and had higher prevalence of tobacco use (52% vs 27%, P = 8E-40), HIV (20% vs 2%, P = 1E-53), and DM (23% vs 5%, P = 5E-45). Among persons living with HIV (PLWH), PTB cases had higher usage of ART (41% vs 0%) and lower CD4 counts (218 ± 225 vs 467 ± 340, P = 1E-06) than controls. Also, 30% of controls received TB infection (TBI) treatment while on study (190 ± 84 days). The risk-controlled sub-cohort fully controlled for HIV and DM (0%) and was more balanced for age (P = 0.07) and TBI treatment (maximum 30 days in controls) (Table 1).

**Figure 1.**
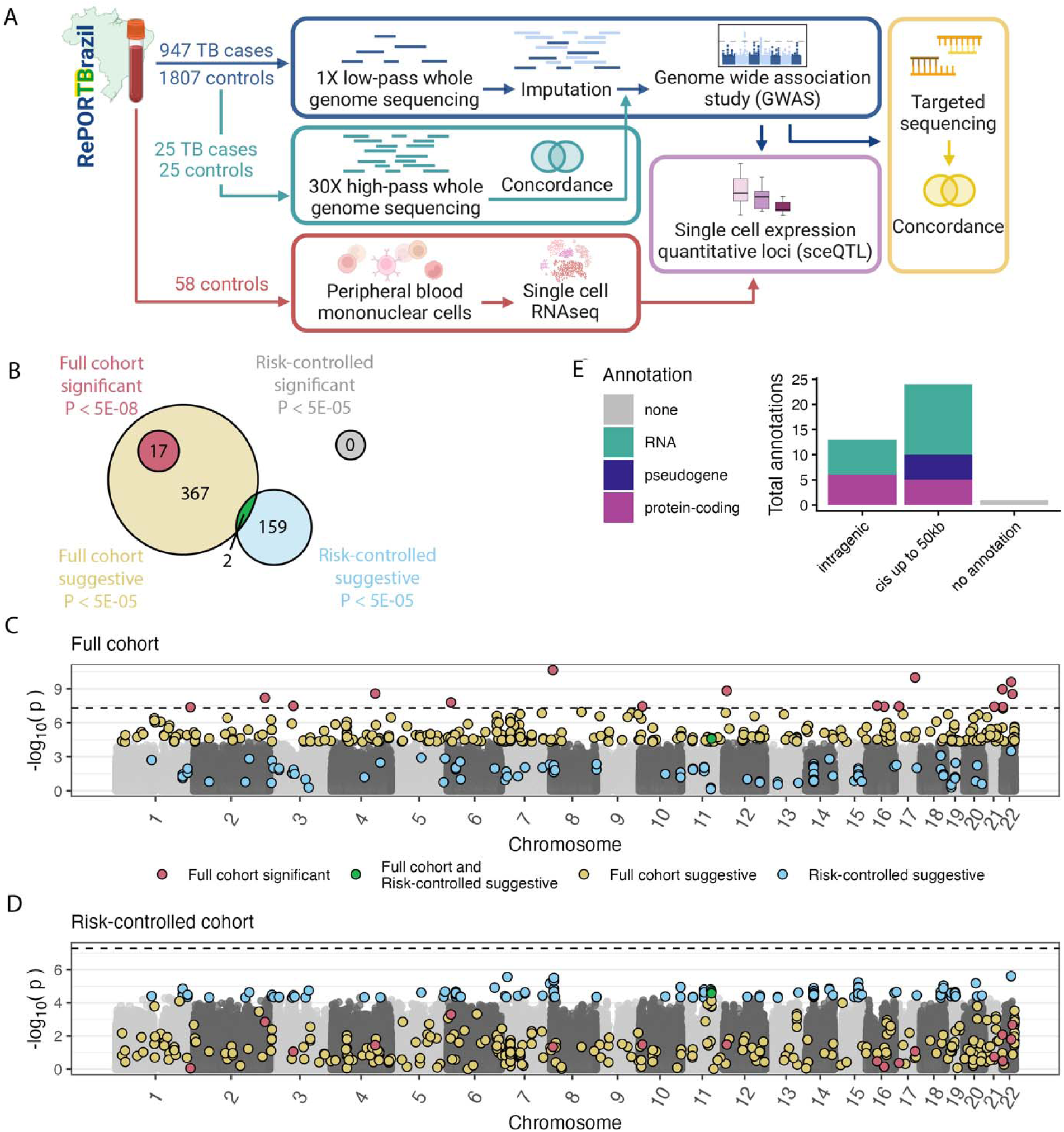
SNPs associated with TB in Brazil GWAS. SNPs at > 5% MAF and verified > 75% concordant in high-pass data are included. (A) Study design and analysis. Additional details can be found in Supplemental Figure 1. Created with biorender.com. (B) In the full cohort, significant SNPs (pink, P < 5E-8) did not overlap with any suggestive SNPs identified in the risk-controlled sub-cohort (blue, P < 5E-5). In total, two SNPs were suggestive in both cohorts (green, P < 5E-5), but no SNPs were significant in the risk-controlled sub-cohort (grey, P < 5E-8). Manhattan plots for the (C) full cohort and (D) risk-controlled sub-cohort in additive mixed effects GWAS model. Concordant genotypes are colored if significant or suggestive in the full and/or risk-controlled cohorts with colors as in (B). For nonsignificant SNPs, alternating odd and even numbered chromosomes are shaded in greys. (E) Gene-level annotations of concordant significant SNPs. Genes are grouped by their closest annotation within a gene or within 50 kb cis of a gene. SNP annotations are colored by protein-coding, pseudogene, RNA (lncRNA, snRNA, snoRNA), and no annotation groups.

**Table 1.**
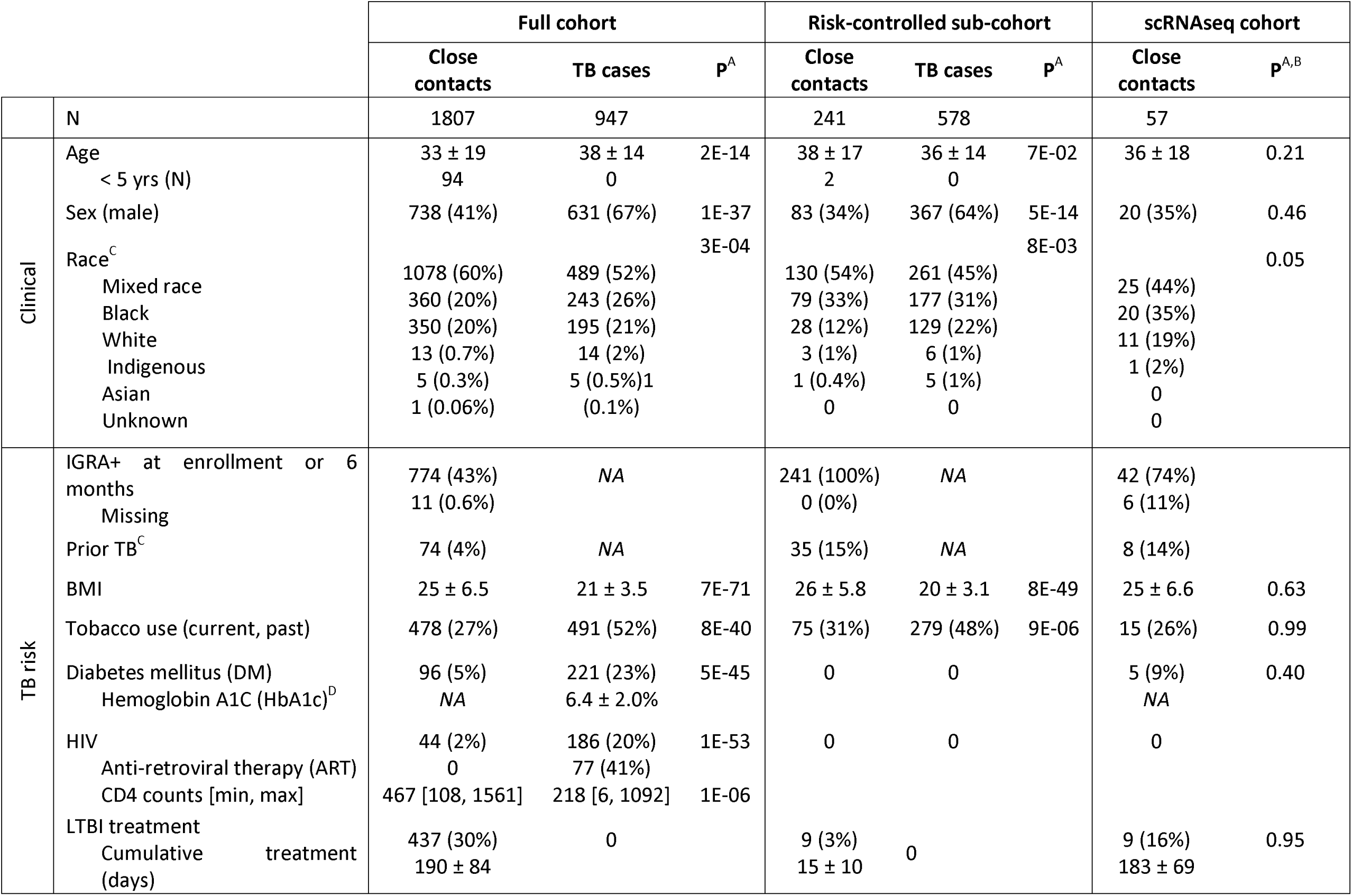
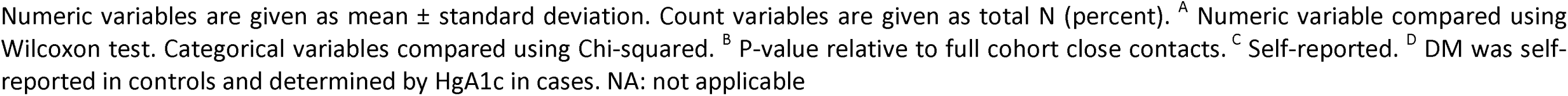
Cohort demographics of Brazil Regional Prospective Observational Research in TB (RePORT) GWAS.

### High-pass genotype confirmation

A random subset of 50 participants (25 PTB cases, 25 controls) was assessed using 30X high-pass whole genome sequencing (hpWGS) to confirm lpWGS imputation results. Selected individuals represented the overall genetic diversity of the cohort as seen in PCA (Supplemental Figure 2A). Of the 12.4 million pass-filter, imputed lpWGS SNPs at 1% minor allele frequency (MAF) (imputation score > 0.6, HWE P > 1 E-6, missingness = 0%, Supplemental Figure 1), the majority (91%) were called in at least 1 participant in the high-pass subset (Supplemental Figure 2B, Supplemental Table 1). Of the high-pass called SNPs, the mean concordance with lpWGS was 99 ± 3.8% and > 99% of SNPs were called in at least 20 participants and ≥ 75% concordance (Supplemental Figure 2C). For GWAS analysis, SNPs were filtered at this concordance, which yielded 6,695,774 SNPs with MAF > 5% (Supplemental Figure 1).

### GWAS SNP-level primary results

Previous PTB GWAS studies commonly corrected for age, sex, and genetic relatedness (ancestry PCs and/or genetic relatedness) (3–6, 8–12, 20, 21). To assess whether to include additional covariates, we tested GWAS additive models on a random sample of 10,000 SNPs for improved fit in the full cohort as measured by reductions in the Akaike information criterion (AIC). We selected a full cohort additive model adjusting for ancestry, sex, age, smoking, HIV, DM, and relatedness (equation (1)). The risk-controlled sub-cohort was modeled for a comparable model removing PTB risk factors (HIV, DM) that were absent in this subset. The full cohort and risk-controlled sub-cohort analyses had an expected quantile-quantile (QQ) trend with the majority of SNPs following a one-to-one ratio and significant SNPs deviating from this trend with low genomic inflation factors (0.976 and 0.998, respectively) (Supplemental Figure 3).

In the full cohort additive model, 17 low-pass, concordant SNPs were significantly associated with PTB disease (P < 5E-08), and an additional 367 SNPs were suggestive (P < 5E-05). The risk-controlled sub-cohort had 161 suggestive concordant SNPs, two of which overlapped with full cohort suggestive SNPs (Figure 1B, Supplemental Table 2). The lack of replication is likely due to limited power in the risk-controlled sub-cohort with only 0.5 to 2.3% power to detect genome-wide significance and 12 to 27% power to detect suggestive significance for the full cohort significant SNPs at their measured MAFs and odds ratios (logistic additive model). Significant SNPs were spread across the genome, including 12 different chromosomes (Figure 1, C and D), and had high-pass concordance of 84 ± 5.5% (Supplemental Figure 2D). Concordance did not differ between cases and controls with 13 to 23 controls and 15 to 23 cases concordant with high-pass WGS for each significant SNP (Supplemental Figure 2E).

### Heritability

Utilizing 488,851 concordant, LD-filtered SNPs across the lpWGS data set and a subset of 1002 unrelated individuals, we estimated heritability by Genome-wide Complex Trait Analysis (GCTA). GCTA found that 40.8 ± 29.3 s.e. % of the variance in PTB disease in this cohort can be explained by genetic factors when corrected for ancestry, age, and sex (Supplemental Table 3). Given the low rate of PTB in the population relative to participants used in the GCTA analysis (0.06 (22) vs 42%), the scaled heritability was estimated to be 13.6 ± 9.8%. When PTB risk covariates were also incorporated (smoking, HIV, DM), heritability estimates were higher at 68.1 ± 30.5% overall and 22.7 ± 10.2% scaled for population prevalence. Together, these data suggest PTB progression in Brazil is partially genetically regulated with high heritability.

### Impact of clinical covariate correction

To assess the impact of PTB risk co-variate inclusion in our study design, we performed analyses with a “standard” GWAS additive model including age, sex, ancestry PCs, and genetic relatedness as well as targeted PTB models lacking one of the PTB risk covariates included in the final full cohort additive model (HIV, DM, smoking). While P-values were correlated between the model with all PTB risk covariates and the “standard” model with no PTB risk covariates (Pearson R = 0.82, P < 2.2E-16), the “standard” model identified only 4 significant SNPs, which fully overlapped with the full model’s results (P < 5E-08, Figure 2A). Thus, accounting for PTB risk factors yielded 13 more significant SNPs without loss of any signal found with a “standard” GWAS model. Among the 17 concordant SNPs significant in the full cohort final additive model, the majority were detected when at least 2 out of 3 PTB risks were accounted for (Figure 2B). Notably, the addition of HIV and/or DM resulted in the greatest increases in significant SNPs detected. Smoking had less impact on significant SNP detection, though 2 SNPs were identified only when all three risk covariates were included. Thus, correction for PTB risk covariates improved model fit and retained statistical power to detect associations with PTB in the full cohort (Figure 2). In addition, the majority of SNPs remained significant despite reductions in power after excluding controls with other potentially confounding factors including LTBI prophylaxis treatment (437 excluded, 11 SNPs significant) or self-reported prior TB (74 excluded, 16 SNPs significant). The remaining SNPs were suggestive (P < 5E-6, Supplemental Table 4), indicating robust genetic associations with PTB.

**Figure 2.**
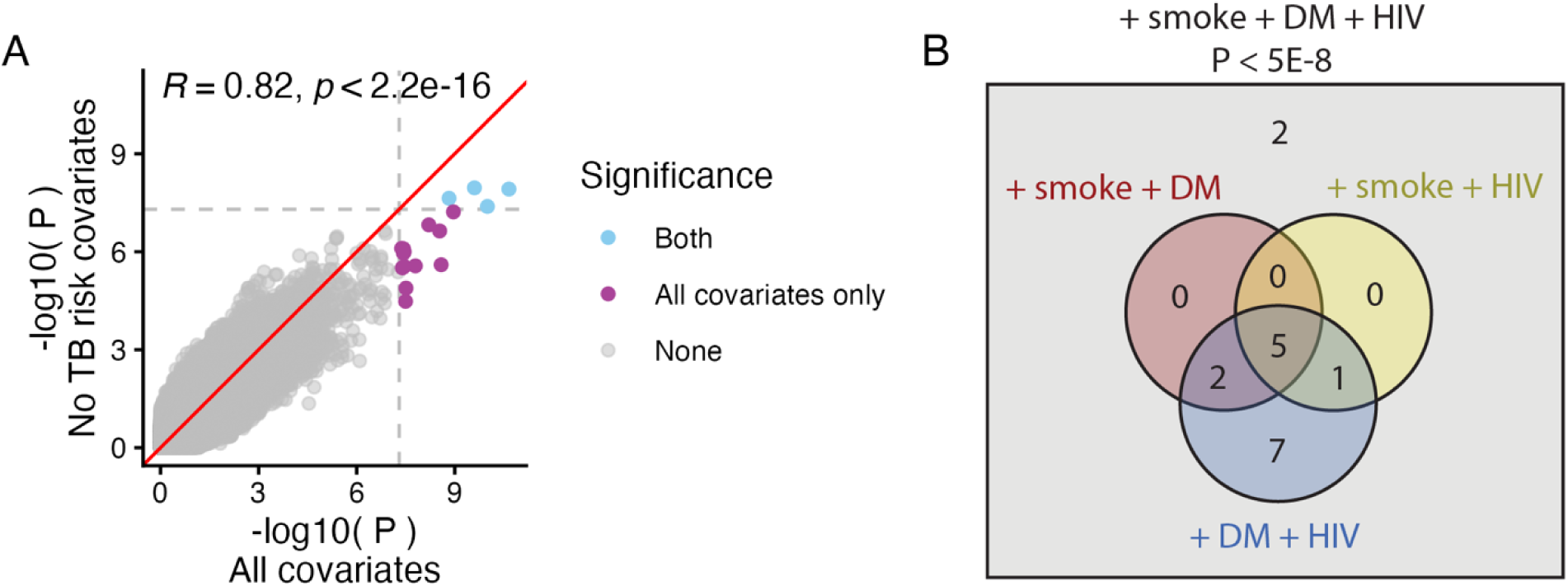
GWAS model impact of correcting for TB risk covariates. The primary full cohort model for TB progressions corrected for ancestry PCs, coefficient of relatedness, age, sex, smoking, HIV, and DM (all covariates). A second, simplified model without TB risk covariates (smoking, HIV, DM) was then applied to the same data. Only SNPs > 5% MAF and > 75% concordance in the high-pass data are shown. (A) Pearson correlation of additive mixed effects P-values for all SNPs in the two models. The red line indicates a 1:1 fit. Dashed lines and colored points indicate model significance (P < 5E-8), and there were no SNPs significant in only the no TB risk covariate model. (B) Impacts of individual TB risk covariates. Leave-one-out models were applied to the 17 SNPs concordant in high-pass and significant in the full cohort model with all covariates (grey box). Significant SNPs in each model lacking one TB risk covariates are totaled in the Venn diagram with 2 SNPs outside the Venn, indicating significance only when all three TB risk covariates are included in the model.

### SNP functional and genomic annotation

The majority of significant, concordant SNPs (16 of 17) were annotated intragenic or within 50 kb cis of genes, including protein-coding, pseudogenes, and RNA genes (lncRNA, snRNA, snoRNA) (Figure 1E, Supplemental Table 4). In total, 6 significant concordant SNPs mapped to 6 protein-coding gene transcripts and 5 SNPs were 50 kb cis to 14 protein-coding genes (Table 2, Figure 3). Among these 20 genes, there was no gene set enrichment within the Hallmark, canonical pathways, or gene ontology databases (FDR < 0.3, overlap > 1, Supplemental Table 5). None of the protein-coding concordant SNPs were significant predictors of a sex, age, smoking, HIV, or DM (Supplemental Table 2), further supporting their association with PTB after correction for these clinical covariates. Further analyses focus on the nine protein-coding annotated SNPs to enrich for potentially causative relationships with PTB disease.

**Figure 3.**
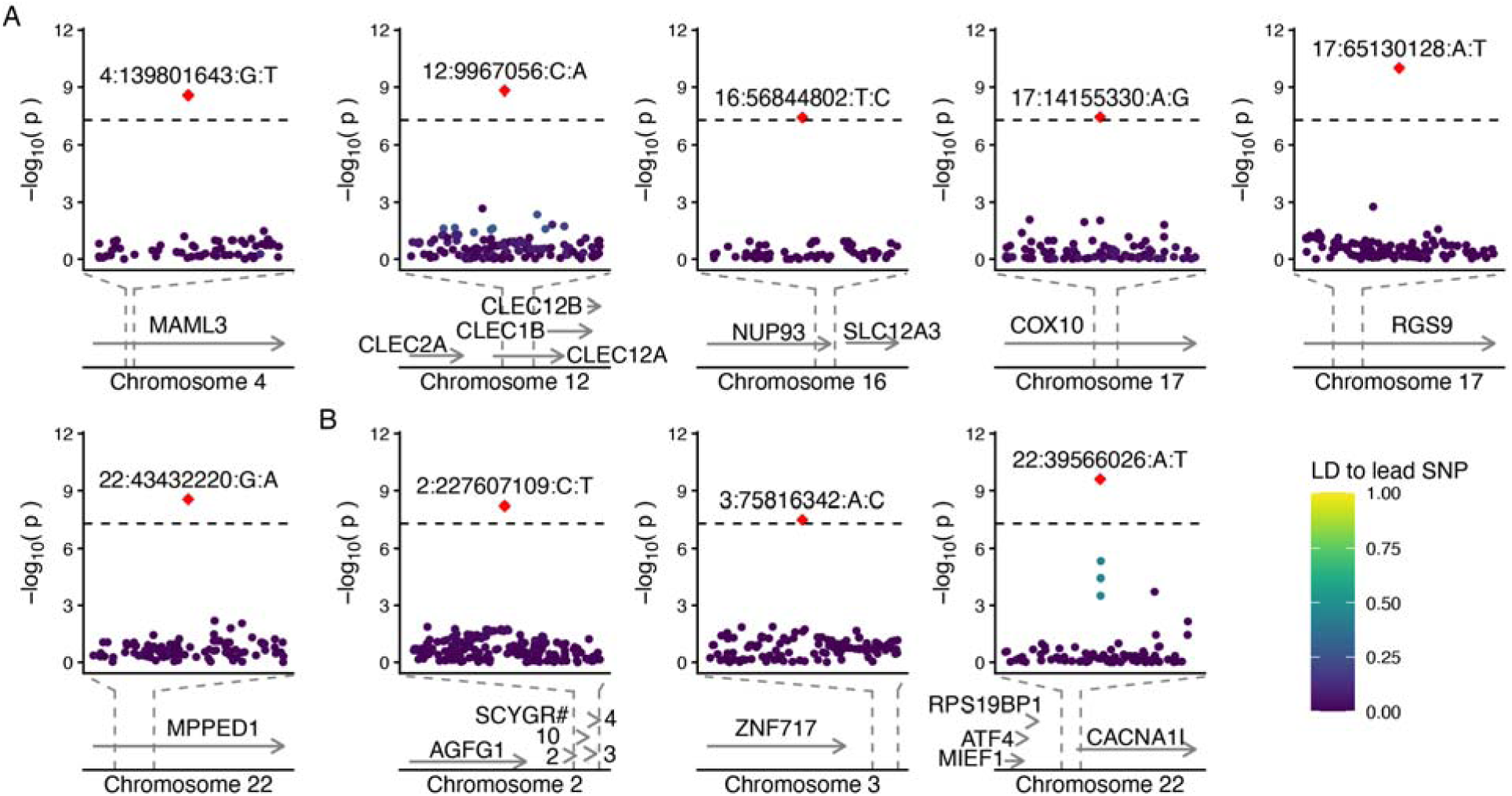
Genomic regions of significant protein-coding SNPs in the full cohort. SNPs significant for TB progression in the full cohort as well as > 75% concordant with high-pass sequencing were selected. SNPs were annotated to protein-coding genes (A) intragenic and (B) cis up to 50 kb. Manhattan plots with concordant SNPs at > 1% MAF and +/- 10 kb from these SNPs are shown with color indicating linkage disequilibrium (LD R) with lead SNPs, which are highlighted as red diamonds. Gene annotations are mapped below with gene boundaries and direction indicated by arrows. Dashed lines indicate the region within the gene map for which GWAS significance is plotted.

**Table 2.**
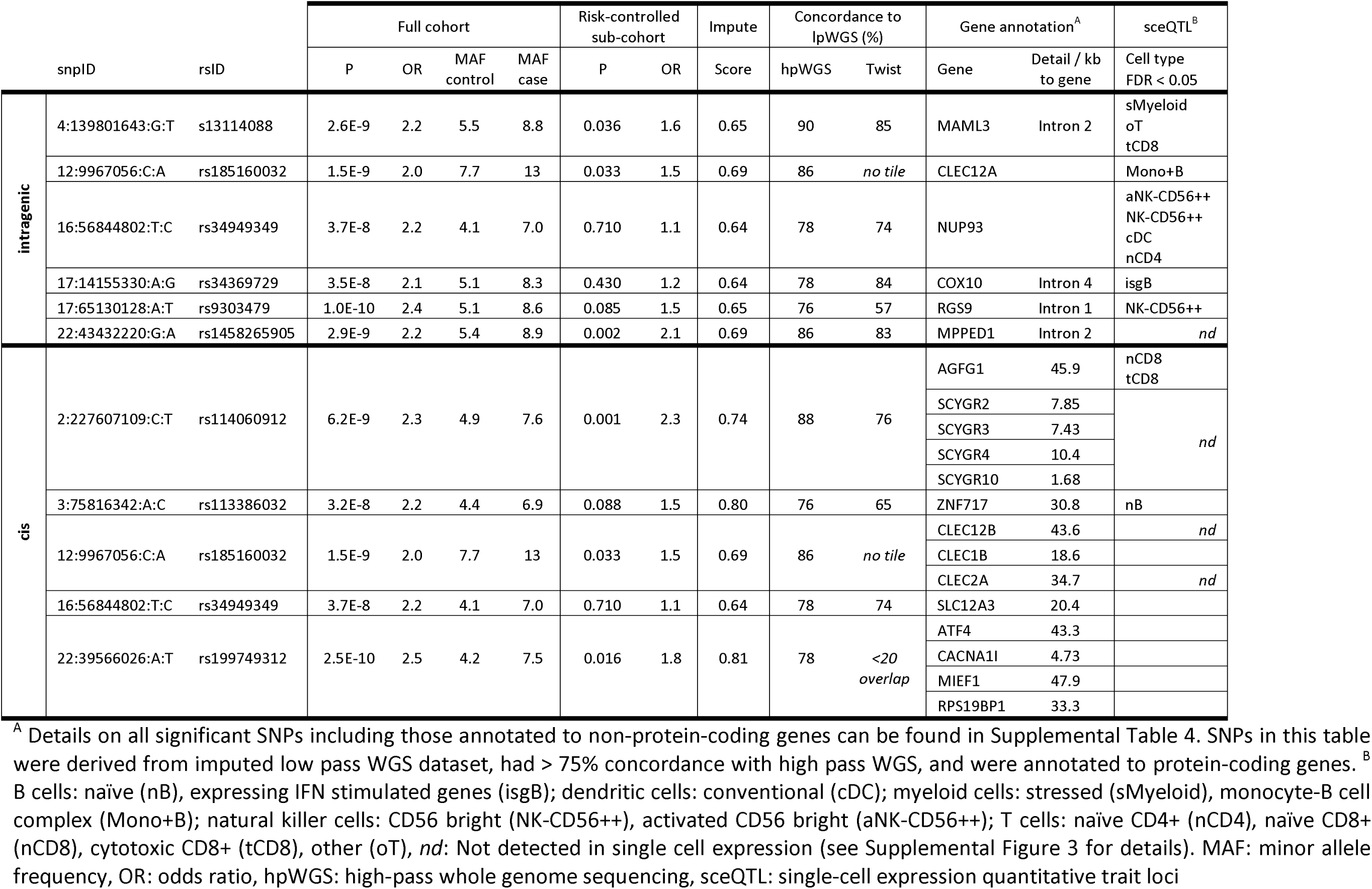
GWAS-level significant SNPs annotated to protein-coding genes.

We next mapped the significant SNPs to assess their genomic and functional features. We assessed lower frequency SNPs (MAF > 1%) within 10 kb of significant concordant GWAS SNPs for linkage disequilibrium (LD R^2^) with the lead SNP (Supplemental Table 2). Significant SNPs did not display clustering of co-inherited, suggestive SNPs due to limited concordant calls and/or low LD in the region (Figure 3).

### Global ancestry estimation and relationship to PTB

ADMIXTURE maximum likelihood analysis was performed against 1000 Genomes to determine global ancestry proportions for RePORT-Brazil participants (Supplemental Figure 4, A and B). Overall, participants most closely resembled the 1000 Genomes Admixed American Super Population with the highest average ancestry proportions of 37 ± 13% for Cluster 5, 32 ± 20% for Cluster 2, and 29 ± 20% for Cluster 4 (Supplemental Figure 4C). In 1000 Genomes, these ADMIXTURE clusters had the highest proportions in European, Admixed American, and African Super Populations, respectively. However, significant variation existed both within the 1000 Genomes Super Populations and within RePORT participants. Utilizing the same logistic linear regression with PTB risk factor adjustment as the GWAS, we found that the proportion of Cluster 3 was moderately associated with PTB progression (P = 0.001, OR = 5.7E5). All other clusters were not significantly associated with PTB (P > 0.4).

### Single-cell eQTL analysis

To assess whether significant concordant SNPs were associated with cis gene expression, we examined a PBMC single-cell RNAseq dataset from the same cohort. Overall, the single-cell subset did not significantly differ from the full cohort controls (Table 1). In total, 57 participants were present in both datasets; this subset was 36 ± 18 years old, 34% male (N = 20), and had similar PTB risk factors as the larger cohort including 25 ± 6.6 BMI, 10% DM (N = 6), 26% tobacco users (N = 15), and no persons living with HIV (PLWH).

SNPs significantly associated with PTB disease (P < 5e-08), concordant in high-pass data (> 75%), and annotated to protein-coding genes (N = 9 SNPs) were modeled against expression of their annotated genes. In total, 8 GWAS-significant concordant SNPs were mapped to 13 protein-coding genes expressed in at least one PBMC cell type in single-cell RNA-seq. CLEC12B, CLEC2A, and MPPED1 as well as SCYGR2, 3, 4, and 10 were not expressed in any cell type; thus, they were not modeled in sceQTL analysis. Among the 32 annotated cell types (Supplemental Figure 5A), all expressed at least one gene of interest, and genes of interest were generally expressed in the majority of cell types with a wide range of expression levels (Supplemental Figure 5B). The exceptions to this were SLC12A3, where was expressed in only 6 B and myeloid cell types, and CACNA1I and CLEC1B, which were expressed in 12 or 14 cell types across all major groups, respectively. In comparison, 18 of 20 protein-coding GWAS-significant genes, including three genes not found in PBMCs here (CLEC12B, MPPED1, SCYGR4), were expressed in tissues relevant to PTB in the Genotype-Tissue Expression (GTEx) Portal, namely lung, liver, or spleen (Supplemental Figure 5C).

Overall, 13 unique SNP-gene-cell additive models were significant eQTLs (FDR < 0.05, Supplemental Table 6). These represented 7 SNPs, 7 genes, and 11 cell types including B, T, myeloid, dendritic, and natural killer cells (Figure 4A). Specifically, SNPs associated with a higher risk of PTB in the clinical GWAS were associated with higher ZNF717 (zinc finger protein), COX10 (cytochrome c oxidase), or CLEC12A (C-type lectin domain) expression in B cell subpopulations (Figure 4B). MAML3 (mastermind-like transcriptional coactivator) expression was associated with PTB risk alleles in both directions with higher expression in T cells (Figure 4C) but lower expression in myeloid cells (Figure 4B). AGFG1 (nucleoporin-like) expression also differed for risk alleles in different T-cell subpopulations (Figure 4C), while NUP93 (nucleoporin) had consistently lower expression associated with risk alleles in T, dendritic, and natural killer cells (Figure 4, C-E). Finally, higher RGS9 (G protein signaling regulator) expression was associated with PTB risk alleles in natural killer cells. Co-localization of GWAS and sceQTL signal was limited (Supplemental Figure 6), likely as a result of limited coverage and LD structure after strict imputation, quality, and high-pass concordance filtering of the data.

**Figure 4.**
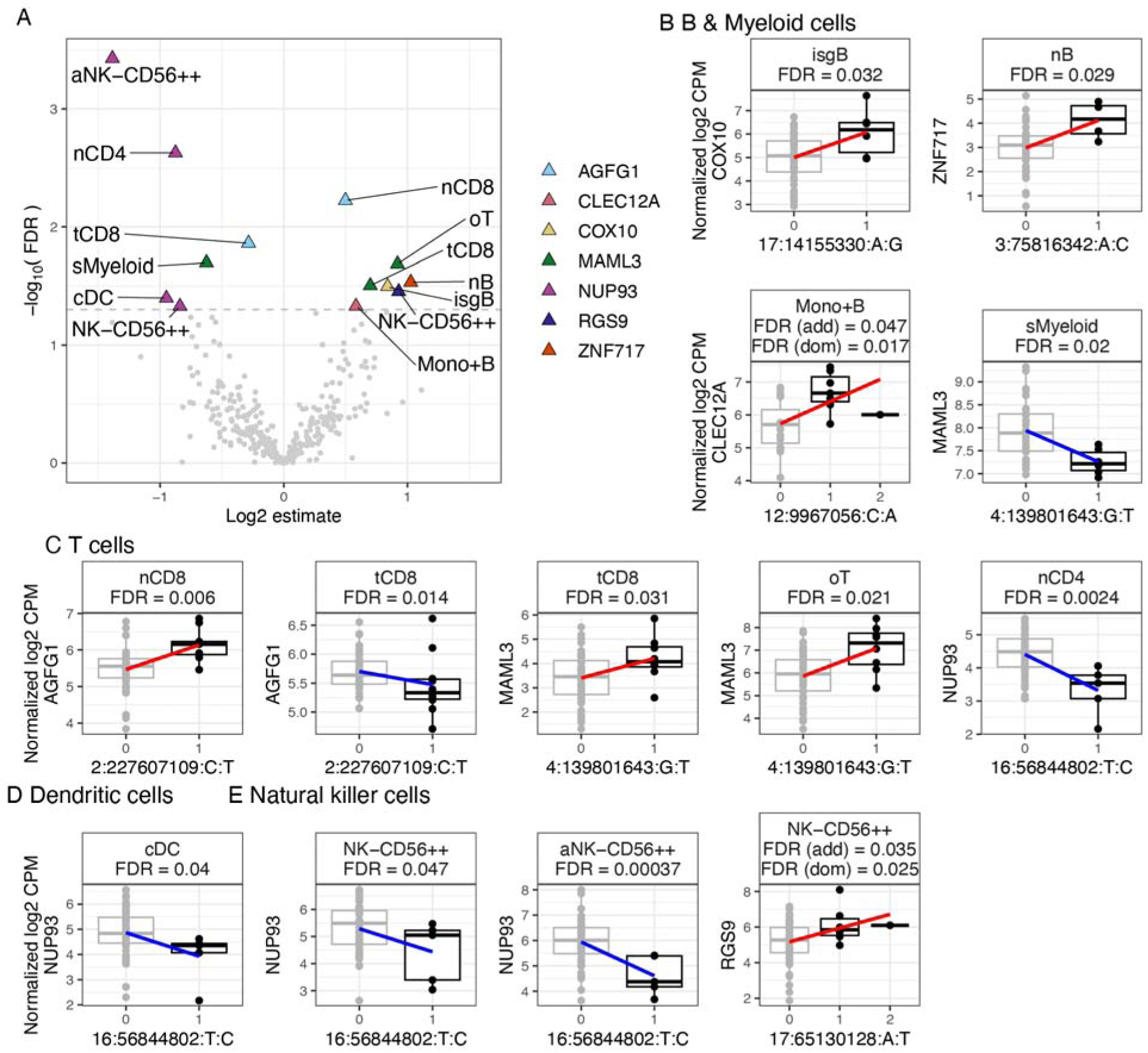
Single cell cis-expression quantitative loci (sceQTL) among TB GWAS significant SNPs. GWAS significant concordant SNPs (P < 5E-8) were mapped 50 kb cis to protein-coding genes that were expressed in single-cell data. Gene expression was modeled against corresponding mapped SNPs for individuals in both data sets (N = 57). (A) Volcano plot of sceQTLs. Color indicates the cis-gene and labels note the single-cell population for significant sceQTLs (FDR < 0.05). SceQTLs present in (B) B and myeloid cells expressing COX10, ZNF717, CLEC12A, or MAML3, (C) T cell populations expressing AGFG1, MAML3, or NUP93, (D) dendritic cells expressing NUP93, and (E) natural killer (NK) cells expressing NUP93 or RGS9. Pseudobulk normalized counts per million (CPM) per donor are plotted in boxplots for sceQTLs. Box plots show the median (center line), interquartile range (IQR, box), and whiskers extending to 1.5X IQR. Genotypes are represented as the number of alternate alleles (0: grey, 1 or 2: black) with SNPs identified by chromosome:position:reference:alternate. Trend lines represent linear models with positive (red) and negative (blue) effects of the alternate allele. Significance is given for the additive mixed effects model (add) and if homozygous alternate individuals are present, also for the dominant model (dom). Cell populations include prefixes for activated (a), conventional (c), cytotoxic (t), IFN stimulated gene+ (isg), naïve (n), other (o), and stressed (s). Full cell population names can also be found in Supplemental Table 6 and Supplemental Figure 5.

### Triple verification by targeted probe-based sequencing

In total, 15 of 17 significant, high-pass concordant SNPs were successfully sequenced via custom probe-based assays. One region (12:9967056:C:A) failed probe design and one region (22:39566026:A:T) yielded < 20 overlapping, pass-filter genotype calls for comparison. Of the 15 Twist called SNPs, 10 were triple verified at > 75% concordance and > 20 calls for lpWGS, hpWGS, and Twist (Table 2, Supplemental Figure 2F). An additional two SNPs were near the 75% cutoff at 72-74% concordance. Concordance between lpWGS, hpWGS, and Twist was correlated (Pearson R > 0.7, P < 0.004), thus supporting the use of either verification method for imputed lpWGS (Supplemental Figure 2G).

### Comparisons to databases and prior GWAS

Of the 17 significant SNPs, six were present on at least one genotype array with the most overlap found for the LIMAArray designed for a Peruvian population (3) (Supplemental Table 7). Despite some overlap, only one SNP (1:238410582:T:C) could be confirmed as non-significant in one study (23) as most prior TB GWAS did not report pre-quality filtering and/or nonsignificant SNPs. Thus, it remains unknown if the continued lack of overlap between TB GWAS is more due to methodology or true genetic differences in populations.

In curatedTBData (24), genes associated with sceQTL were detected in 6 to 9 bulk RNAseq or microarray whole blood studies queried (Supplemental Table 8). Specifically, AGFG1, CLEC12A, and MAML3 were more highly expressed and COX10, NUP93, RGS9, and ZNF717 were more lowly expressed in whole blood from individuals with PTB. These results were directionally consistent and significant (P < 0.05) for 1 to 7 independent studies per gene. In addition, significant sceQTLs were compared to the Database of Immune Cell Expression (DICE (25)). Notably, the intronic RePORT sceQTL for COX10 in IFN-expressing B cells (17:14155330:A:G) was an eQTL in naïve B cells along with 25 additional eQTLs within 10 kb in DICE (Supplemental Table 9). NUP93 (18 eQTL), ZNF717 (7 eQTL), and AGFG1 (3 eQTL) also had DICE eQTLs near RePORT sceQTLs in the same cell types: naïve CD4+ T-cells, naïve B-cells, and naïve CD8+ T-cells, respectively. Finally, MAML3 (6 eQTL) and RGS9 (1 eQTL) had nearby DICE eQTLs in similar cell populations, though not exactly matched (Supplemental Table 9).

## DISCUSSION

We used a genome wide association study (GWAS) to discover genetic variants associated with pulmonary tuberculosis (PTB) in Brazil with four important study design features: inclusion of extended epidemiologic risk factors in case-control definitions, genotyping with imputed low-pass whole genome sequencing (lpWGS) to capture more comprehensive population-specific variants, verification of imputation results by two independent methods, and the assessment of the functional significance of candidate genetic variants with single-cell eQTL (sceQTL) analysis in the same cohort. We discovered several genetic variants associated with PTB with genome-wide level significance and associated function in cell-specific sceQTLs.

Overall, the heritability of PTB in Brazil was estimated to be 40 to 68% (scaled 14 to 23%) depending on inclusion of the TB risk factors HIV, DM, and smoking. This is within the range of estimates from other populations, including Uganda (55% (23)), China (38% (21)), and Peru (21% (3)), which did not correct for TB risks. The variability across studies may reflect differences in genetic background, environmental exposures, epidemiological context, and study design, in addition to technical factors such as genotyping arrays, imputation panels, and analytical approaches. Together, these findings support a substantial host genetic contribution to PTB susceptibility across diverse populations, while also underscoring the importance of context-specific factors that may modulate the observed heritability.

To gain insight into genetic regulation of susceptibility to PTB, we examined high-confidence imputed lpWGS SNPs that were associated with PTB disease, concordant in high-pass WGS (hpWGS), mapped to protein-coding genes, and expressed in a matched single-cell RNA-seq experiment. Notably, two SNPs associated with nucleoporins were significantly associated with PTB and sceQTLs as well as near triple verification by hpWGS (concordance > 75%) and targeted sequencing (concordance > 74%). An intronic SNP was associated with lower expression of the nucleoporin NUP93 in multiple cell types, including naïve CD4+ T cells, and a downstream SNP was associated with higher expression of the nucleoporin-like AGFG1 in multiple T cell populations, including naïve CD8+ T cells. Both genes were also significantly differentially expressed with PTB in whole blood transcriptomes (NUP93 (26, 27), AGFG1 (28, 29)) as well as had multiple directionally congruent eQTLs in DICE (25). Nucleoporins mediate nucleocytoplasmic transport with NUP93 acting as the central scaffold protein within the nucleoporin inner ring complex (30). Depletion of NUP93 results in destabilization of the nuclear pore complex and is associated with reduced innate immune responses like TBK1 activation (31). Changes to TBK1 activation may impact PTB progression as a result of TBK1’s role in autophagy-mediated antimicrobial defense (32). Therefore, multiple lines of independent evidence support that genetic regulation of nucleoporin genes may contribute to PTB risk and progression.

Additionally, an intronic mastermind-like transcriptional coactivator 3 (MAML3) SNP was associated with PTB risk and gene expression in multiple cell populations as well as triple-verified by hpWGS and targeted sequencing. Specifically, the MAML3 intronic SNP was associated with decreased expression of MAML3 in stressed myeloid cells, and a directionally congruent eQTL was present less than 100 bp away for classical monocytes in DICE. In contrast, the same MAML3 SNP was associated with increased expression in other cell types, including CD8+ cytotoxic T cells, which directionally matched prior whole blood bulk transcriptional data where MAML3 was more highly expressed with PTB (29, 30, 33). Despite this SNP’s presence on the Affymetrix LIMAArray (3), it could not be confirmed as it failed MAF and/or HWE filters in the prior Peruvian study, and thus, no P-value was reported. MAML3 is a transcription coactivator involved in positive regulation of transcription, particularly Notch signaling. Notch pathways have been implicated in PTB disease including T cell cytokine production (34), Th1/Th2 balance (35), and apoptosis in response to Mycobacterium tuberculosis (Mtb) antigen (36). Thus, genetic regulation of Notch through MAML3 may contribute to PTB risk.

Moreover, an intronic Regulator of G protein signaling 9 (RGS9) SNP was associated with increased PTB risk and RGS9 gene expression in CD56-bright natural killer (NK) cells. While the canonical role of RGS9 is regulation of neuronal G protein signaling pathways (37, 38), it has been linked to a tissue resident-like phenotype in NK cells (39). In particular, CD56-bright NK cells are characterized by higher interferon-gamma (IFNG), tumor necrosis factor-beta (TNFB), interleukin-10 (IL10), and IL12 production in response to monocyte-derived cytokines when compared to their CD56-dim counterparts (40). Thus, these NK-derived cytokines may provide an important innate immune response to PTB downstream of primary monocyte infection. These data suggest a possible genetic regulator of the tissue resident-like NK phenotype through RGS9, though verification in a larger sample size would be beneficial as the SNP only pass hpWGS verification, not targeted sequencing.

Finally, a SNP downstream of zinc finger protein 717 (ZNF717) was associated with this gene’s expression in naïve B cells. ZNF717 encodes a Kruppel-associated box (KRAB) zinc-finger protein, which binds DNA and has predicted roles in regulation of transcription, but no published experimental studies of its function. However, we found lower ZNF717 expression in whole blood RNAseq associated with PTB in vivo (28) as well as seven previously published naïve B cell eQTLs within 10 kb in DICE (25). Here, the SNP downstream of ZNF717 was associated with increased expression in naïve B cells, which was congruent with DICE eQTLs but not bulk transcriptomics, possibly due to the relatively low contribution of B cells to bulk blood expression. Together, these data suggest that genetic regulation of ZNF717 impacts corresponding gene expression and is associated with increased susceptibility to PTB. Given the cis annotation of this SNP and < 75% concordance with targeted sequencing, additional in vitro work is necessary to verify ZNF717’s potential role in TB. Overall, the combination of GWAS and sceQTL datasets in the same cohort enables rapid screening of GWAS SNPs for those most likely to impact PTB disease through transcriptomic outcomes. The single-cell approach further facilitates detailed identification of the involved cell types as well as directionality and magnitude of effects.

Our study had several limitations. This study aimed to investigate genetic determinants of symptomatic PTB. Thus, controls included TB exposed, LTBI prophylaxis treated, and potentially TB infected individuals as evidenced by IGRA positivity. Therefore, subclinical or asymptomatic TB as well as TST/IGRA conversion were not investigated, though TST/IGRA positivity represents an important risk for PTB disease and has been investigated in several prior GWAS (23, 41, 42). In addition, LTBI treatment may have reduced risk of progressing to symptomatic disease regardless of genetic susceptibility, potentially masking GWAS associations. This may bias results toward the null and should be considered when interpreting findings, particularly as they relate to overlap with other GWAS.

There were also a number of technical limitations in this study. First, although one distinguishing feature of this study was its use of unbiased genome-wide genotyping with lpWGS, imputation discrepancies did exist between lpWGS, hpWGS, and probe-based sequencing, even among SNPs that passed lpWGS imputation quality and minor allele frequency (MAF) filtering. This may indicate genomic regions prone to low quality sequencing and/or imputation that should be excluded in future studies. However, testing with additional populations and reference data sets is necessary to confirm these regions, particularly given the sparsity of Brazilian individuals in current references. Nevertheless, this method yielded millions of SNPs not represented by current or past SNP arrays, and overall, imputed lpWGS genotype calls were highly concordant with hpWGS and probe-based sequencing.

Second, this study cohort displayed diverse ethnic and geographic backgrounds (43) as well as variability in a number of known PTB risk factors. Traditionally, GWAS correct for age, sex, and ancestry as well as often exclude individuals with known risk factors like HIV. Here, we have shown through statistical assessment that exclusion was not necessary, and many factors can be accounted for as model covariates. This preserved sample size, and thus statistical power, which would otherwise be reduced in case-control designs. Specifically, PTB risk factors including HIV, DM, and smoking improved model fit as well as increased PTB signal despite the more complex statistical model. The smaller risk-controlled sub-cohort failed to identify these statistically significant SNPs, likely as a result of reduced power. This underscores the importance of these factors in PTB risk assessment and may contribute to the lack of reproducibility among TB GWAS that did not account for similar risk factors. Third, the sample size of this PTB GWAS did not allow for the detection of rare variants (MAF < 5%) and was under-powered to detect SNPs with small effect sizes.

Fourth, the sceQTL analysis was performed in an unstimulated PBMC dataset, which limits our findings to baseline expression of protein-coding in these cell types. Expression patterns of some genes suggest that mechanisms of action may occur in tissues such as the lung, liver, or spleen, and additional sceQTL effects may only be apparent in response to infection or other stimuli. Also, mechanisms for SNPs impacting non-protein-coding genes represent a rich, herein unexplored avenue to explain the heritability of TB risk. Finally, despite advances in design and methodology, this GWAS failed to identify PTB-associated SNPs of significance described in prior GWAS, even SNPs present on prior SNP arrays. This is a common feature among TB GWAS and may indicate an abundance of population-specific genetic risk factors. Mtb has co-evolved with humans for thousands of years, and there is considerable Mtb strain variability worldwide (44). Thus, population-specific SNPs relevant to PTB disease risk are likely present, and unbiased genotyping across multiple populations will be necessary to identify population-spanning SNPs.

In summary, this PTB GWAS identified 17 loci associated with PTB disease, nine of which were associated with protein-coding genes and seven had potential functional consequences identified through sceQTL analysis. These findings provide new insights into genetic risks of PTB in the highly admixed Brazilian population. While no overlap was found between this and prior TB GWAS, future studies employing lpWGS may yield population-spanning SNPs not previously measured across populations due to the limitations of prior SNP arrays. Thus, this study provides important groundwork for both lpWGS methods and the assessment of genetic risks for PTB.

## METHODS

### Cohort

From 2015 to 2019, culture-confirmed adult pulmonary tuberculosis (PTB) patients were recruited at study clinics in Rio de Janeiro, Manaus, and Salvador as part of the Regional Prospective Observational Research in TB (RePORT) study in Brazil. Full details were reported previously (45). Briefly, PTB cases were confirmed by positive Mycobacterium tuberculosis sputum culture at enrollment (N = 1,060, Supplemental Figure 1). Close contacts of PTB patients were then recruited using cumulative density sampling under the criteria of > 4 hrs/wk of contact with the index PTB cases within the past 6 months (N = 1,839). Controls were defined as close contacts with normal chest radiograph at enrollment and no symptoms for 24 months of follow-up. Interferon-gamma release assays (IGRA) were performed for controls at enrollment and a 6-month follow-up, but negative IGRA or prior TB were not considered a control requirement. Additional radiograph tests and sputum were only performed when contacts presented with symptoms during the course of the study. Close contacts who were asymptomatic with normal chest radiograph at enrollment but progressed to active PTB during this time the two-year study follow-up were classified as progressors (N = 25) and excluded from the GWAS.

### Risk-controlled sub-cohort

Among the 2,754 pass-filter individuals (see below), 819 individuals (241 controls, 578 PTB cases) were included in a risk-controlled sub-cohort in which all were HIV-seronegative and without diabetes mellitus (DM). DM was self-reported for controls and defined by HbA1C at enrollment < 6.5 for cases. In addition, controls were IGRA-positive at enrollment, received < 30 days of any treatment for tuberculosis infection during the course of the study, and had ≥ 12 months of follow-up during which they did not develop active PTB. Treatments included isoniazid or rifampin monotherapy. Sensitivity analyses were performed for significant SNPs (see GWAS statistics) excluding individuals with prior TB (self-reported, N = 74) or LTBI prophylaxis treatment (N = 437).

### Sample collection and DNA library preparation

At enrollment, whole blood was collected in EDTA tubes, aliquoted, and immediately frozen at −80 °C. DNA was extracted from 100 μl of whole blood using the DNeasy Blood and Tissue Kit (Qiagen) following the manufacturer’s protocol with an additional final centrifugation to maximize DNA recovery. DNA was quantified using Qubit Fluorometer (Thermo Fisher) and aliquoted in 96 well plates to 10 ul at 100 ng/ul. Libraries were prepared using the Twist Biosciences kit (P/N: 104207) following a miniaturized version of the manufacturers protocol and quantified using Qubit. Pool quality was assessed using the Agilent Bioanalyzer and quantified using a qPCR-based method with the KAPA Library Quantification Kit (P/N: KK4873) and the QuantStudio 12K instrument. Prepared DNA library pools were pooled for sequencing in equimolar ratios based on qPCR.

### Low-pass sequencing and genotype calls

In total, 2791 participants were sequenced using Illumina low-pass whole genome sequencing (lpWGS) at 1-2X coverage at Vanderbilt University Medical Center Technologies for Advanced Genomics (VANTAGE). DNA library pools were subjected to cluster generation using the NovaSeq 6000 System, following the manufacturer’s protocols. Sequencing was performing using 150 bp paired-end reads on the NovaSeq 6000 platform targeting 10 million reads per sample (1-2X genomic coverage). Raw sequencing data (FASTQ) was subjected to quality control analysis, including read quality assessment. Real Time Analysis Software (RTA) and NovaSeq Control Software (NCS) (Illumina) were used for base calling. MultiQC (v1.7; Illumina) was used for data quality assessments. Single nucleotide polymorphisms (SNPs) were called using Illumina DRAGEN Germline v4.0.3 with default low-pass parameters in Illumina BaseSpace. Genotypes were then imputed using Illumina DRAGEN Imputation v4.0.3, which implemented GLIMPSE (13) against the 1000 Genomes Project Phase 3 (46) IRPv1 autosomal SNP reference panel (N = 2,489). Imputation resulted in 49.5 million autosomal SNPs, which were filtered to 31.1 million (63%) with imputation score > 0.6. Genotypes were further processed using PLINK2 (47, 48) and R v4.4.0 (49). SNPs on X and Y chromosomes were not included in the analysis. In addition, 3 samples (2 cases, 1 contact) failed quality controls prior to imputation and were not included in analyses (Supplemental Figure 1).

### Ancestry and kinship

Imputed genotypes were filtered by minor allele frequency (MAF) > 1% and sliding window linkage disequilibrium (LD) with a 500 kb window, 1 kb slide, and maximum r^2^ of 0.2 using PLINK2. With the resulting 12.6 million genotypes, identity by descent was calculated using the Robust KING method (50) in SNPRelate (51) and ancestry principal components (PCs) were estimated using PC-AiR (50, 52) in GENESIS (53). Kinship was then calculated using PC-Relate (54) with 2 ancestry PCs and a training set of the 1,554 unrelated individuals defined by PC-AiR. Sample pairs with high relatedness (kinship > 0.4) were further assessed and determined to be twins (N = 5 pairs), siblings (N = 1), duplicate samples (N = 5), or unrelated/unknown (N = 2). The first chronological sample was retained for duplicates, and high-kinship unrelated/unknown pairs were removed from analysis. This resulted in 2,754 individuals for GWAS analyses (1,807 controls, 947 PTB cases).

Pass-filter samples were compared to 1000 Genomes Project Phase 3 (46) (N = 2,548) using principal component analysis (PCA). RePORT imputed genotypes were filtered to MAF > 5%, Hardy-Weinberg Equilibrium (HWE) P > 1E-6, 0% missingness, and ≥ 75% concordance with high-pass WGS (see Genotype validation by high-pass sequencing). Overlapping filtered RePORT and 1000 Genomes genotypes were then filtered to remove AT and CG SNPs, which are more prone to sequencing error, and LD filtered using a sliding window of 50 bp, 10 bp slide, and r^2^ threshold of 0.1. Finally, genotypes were filtered to MAF > 10% in the combined dataset. Using these 139,855 SNPs, PCs were calculated using PLINK2, and RePORT participant self-reported race was compared to 1000 Genomes Super Populations. RePORT participants reported race under a multiple-choice single response question including Pardo (mixed race), Negro (Black), Branco (White), Amarelo Asiático (Asian), or Índio (Indigenous). One participant failed to respond to this question but was retaining in analyses since ancestry PCs were used in the GWAS in lieu of self-reported race. In addition, local ancestry proportions were estimated using maximum likelihood in ADMIXTURE (55) with K = 5 aimed at representing the 5 Super Populations in 1000 Genomes.

### Genotype validation by high-pass sequencing

To validate imputed genotypes, a random subset of individuals (25 controls, 25 cases) was re-sequenced with 30X genome coverage at VANTAGE. DNA library pools were subjected to cluster generation using the NovaSeq X Plus System, following the manufacturer’s protocols. Sequencing was performed using 150 bp paired-end sequencing on the NovaSeq X Plus platform targeting 350 - 400 million reads per sample (30X genomic coverage). Raw sequencing data (FASTQ) was quality assessed in FastQC, and bases were called as for low-pass sequencing. Sequences were trimmed using fastp (56) (5-prime trimming = 10 bp, adapters = Illumina universal) and aligned with bwa-mem2 (57) against the human genome GRCh38 formatted for the Genome Analysis Toolkit (gatk (58)). SNPs were called following GATK best practices for germline short variant discovery (59). Briefly, alignments were filtered for primary alignments with a minimum mapping quality score of 20, and duplicates were marked. Base quality score recalibration (BQSR) was applied against dbSNP, Mills gold-standard indels, and Broad known indels. SNPs were called at genomic positions present in the low-pass data set using HaplotypeCaller, and merged using joint genotyping. Multi-allelic SNPs were split, and calls were converted to PLINK2 (48). This resulted in 11.3 million (91%) SNPs overlapping with pass-filter low-pass data at 1% MAF. Concordance between low- and high-pass genotypes was calculated as the percent identical genotype calls. SNPs with concordance ≥ 75% and at least 20 overlapping calls progressed to GWAS analysis.

### GWAS model fitting

From the 12.6 million genotypes used for kinship, 10,000 random positions were selected for model fitting. Mixed effects logistic regression was performed to test genotype associations with PTB cases (1) vs close-contact controls (0) (Supplemental Table 2). Participants predominantly identified as mixed race (57%), black (22%), or white (20%), which most closely corresponded to 1000 Genomes (46) admixed American, African, and European Super Population ancestry groups, respectively, though significant admixing was apparent (Supplemental Figure 4, A and B). ADMIXTURE analysis revealed that RePORT-Brazil participants most closely resembled the 1000 Genomes Admixed American Super Population (Supplemental Figure 4C).

Ancestry PCs were progressively added to the base GWAS additive model and assessed for total variation explained as well as model fit by Akaike Information Criterion (AIC). A covariate was retained if it significantly improved model fit in its univariate model and its removal worsened model fit in its leave-one-out model (|ΔAIC| > 8). The first two ancestry PCs were included in the final additive model as they explained the most variation (PC ≥ 1%) (Supplemental Figure 4D) and improved model fit for the majority of SNPs as indicated by reduced AIC compared to models with one fewer PCs (Supplemental Figure 4E). Next, the coefficient of relatedness (2 x kinship) was added to the 2 PC logistic additive model as a pairwise covariance random effect and tested with GMMAT (60). While relatedness did not improve model fit (Supplemental Figure 4F), the cohort contained 1,723 potential first-degree relationships (kinship > 0.2), including five confirmed twin pairs (Supplemental Figure 4G). Thus, the coefficient of relatedness was retained in the additive model to account for this cohort structure.

Finally, additional clinical covariates age and sex as well as PTB risk covariates smoking, HIV status, and diabetes mellitus (DM) (self-reported for controls, enrollment HbA1C > 6.5 for cases) were assessed. Each covariate was assessed in univariate additive models (e.g. 2PC relatedness model + one covariate, Supplemental Figure 4H) and leave-one-out additive models (e.g. 2 PC relatedness model + all covariates except one, Supplemental Figure 4I). Smoking was classified as past/current vs never as there were no significant differences between past and current smokers in a univariate additive model (Supplemental Table 2, P > 0.3). All covariates except age significantly improved model fit when added in univariate models and worsened model fit when removed in leave-one-out models (Supplemental Table 2). Age initially worsened model fit when added to the simpler model but had no significant effect when removed from the more complex model (|ΔAIC| < 8). Thus, age was retained in the final model to facilitate comparisons to previous GWAS studies that commonly corrected for age. This resulted in the full cohort additive model

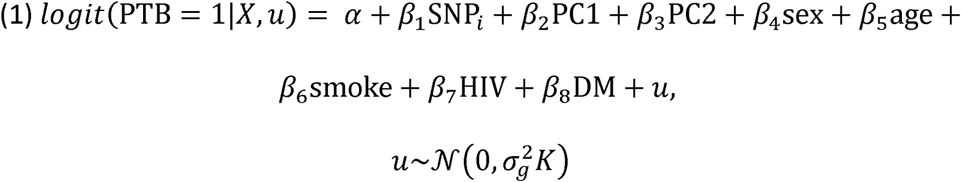

where logit(P(TB=1∣X, u)) is the log-odds of PTB disease given the covariates X and random effects u, α is the intercept, β are the coefficients of main effects (SNP) and covariates (ancestry PCs, clinical covariates, PTB risk covariates), and u is the random effects with covariance structure determined by genetic relatedness (K). The risk-controlled sub-cohort was modeled for a comparable model removing PTB risk covariates (HIV, DM) that were absent in this subset. Global ancestry proportions were modeled in a similar model omitting ancestry PCs and replacing the SNP effect with proportions for clusters 2 through 5.

### GWAS statistics

The full dataset imputed 49.5 million genotypes were filtered for imputation score > 0.6, high-pass concordance ≥ 75% with at least 20 calls, MAF > 5%, HWE P > 1E-6 (calculated on controls only), and 0% missingness. This resulted in 6,695,774 genotype positions for analysis. The full cohort was modeled for additive SNP effects with adjustment for ancestry, relatedness, clinical, and PTB risk covariates as in equation (1), and the risk-controlled sub-cohort with this model without HIV and DM as person living with HIV and/or DM were excluded from this analysis. Both cohorts were additionally analyzed with a simplified additive model without all three PTB risk covariates (smoking, HIV, DM) to compare with previous GWAS studies.

The rationale for the full cohort analysis was to maximize sample size and potential statistical power while adjusting for covariates that could decrease precision without fully addressing confounding bias. The rationale for the risk-controlled sub-cohort was to optimize case and control selection to exclude variability introduced by risk factors known to be associated with PTB disease (e.g. HIV, DM). The risk-controlled design helped improve statistical efficiency for estimating the causal effect within and across strata of balanced characteristics (relying on discordant pairs to be informative) and more powerfully limited the confounding bias for important a priori-identified confounders. Though the risk-controlled design lent itself to estimating a causal effect less prone to bias and more likely to be internally valid, statistical power was reduced relative to the full cohort sample size. The merits of the full cohort analysis (improved power to detect small effects) and risk-controlled design (improved internal validity to detect unbiased causal effects) are, therefore, complementary instead of redundant.

All genotypes were scored using GMMAT (glmm.score) (60) in the full and risk-controlled cohorts separately. Genotypes with a score P < 1E-4 were then fit in the full GLMMkin additive model. In addition, all full cohort selected genotypes were also run in the risk-controlled sub-cohort. Significant SNPs were defined at P < 5E-8 and suggestive SNPs at P < 5E-5. Lower MAF genotypes (1 - 5%) were then selected within 10 kb of significant SNPs and tested with the same models described above as well as assessed for linkage disequilibrium (LD) with significant SNPs in the region to create locus-zoom plots. Significant SNPs were also tested for associations with clinical and PTB risk covariates by modeling sex, age, smoking, HIV, or DM as outcomes in place of PTB status while retaining all other covariates. Finally, significant SNPs were cis-mapped to genes within 50 kb. Gene enrichment was performed against Broad MSigDB (61) Hallmark, canonical pathways, and gene ontology using iterative hypergeometric enrichment in SEARchways (62) to account for SNPs with multiple gene annotations.

The overall heritability of PTB progression was calculated using Genome-wide Complex Trait Analysis (GCTA (63)) with a genetic relationship matrix (GRM) and restricted maximum likelihood (REML (64)) calculated from 488,851 concordant, LD-filtered SNPs (r^2^ > 0.2, 50 bp window, 5 bp slide) in 1002 unrelated individuals from the full cohort (584 controls, 413 cases, GRM < 0.05). Prevalence was estimated as the mean PTB prevalence during the recruitment period (2025–2019) in RePORT participating Brazilian states (Amazonas, Bahia, Rio de Janeiro) according to the Pan American Health Organization (57.07 cases per 100,000 population) (22). Additive models were corrected for covariates as in the full cohort (ancestry PCs, sex, age, smoking, HIV, DM) as well as without PTB risk covariates for comparison.

### Genotype validation by targeted sequencing and database overlap

A subset of 95 individuals (73 controls, 22 cases) was selected for targeted sequencing with Twist probe-capture enrichment. In total, probes tiling regions +/- 25 kb around 16 of 17 significant, hpWGS concordant SNP were successfully designed (excluding 12:9967056:C:A). DNA was extracted from fresh EDTA aliquots via the same protocol as lpWGS. Twist libraries were constructed and pooled using the Twist Biosciences kit (P/N: 104207) following a modified, miniaturized version of the manufacturers’ protocol for the Biomek i7 liquid handler. The pool was captured using custom probes and a modified protocol to support high-multiplexing with probes designed to capture smaller regions. The library quality was assessed using the Agilent Bioanalyzer and quantified using a qPCR-based method with the KAPA Library Quantification Kit (P/N: KK4873) and the QuantStudio 12K instrument.

Prepared libraries were pooled in equimolar ratios, and the resulting pool was subjected to cluster generation using the NovaSeq XP System, following the manufacturer’s protocols. Sequencing was performed using 150 bp paired-end reads targeting 1-2M reads per sample. Raw data were quality-controlled and base-called as with low-pass WGS. Targeted sequencing data was then processed and variants were called using the same GATK germline pipeline as high-pass WGS. Concordance between targeted genotypes and low- or high-pass data was calculated as the percent identical genotype calls. SNPs with concordance ≥ 75% and at least 20 overlapping calls for all three pairwise comparisons were considered “triple concordant”.

Significant GWAS SNPs were also verified by comparing to SNP manifests for 19 arrays used in prior TB GWAS studies. UCSC LiftOver (65) was used to convert this study’s hg38 genomic locations to older genome versions (hg37, hg36, hg35) for comparisons. Only 7 of the 17 significant concordant SNPs were present on one or more arrays. The most overlap was found with the custom LIMAArray (6 SNPs), which was designed for a Peruvian population (3).

### Single-cell RNA sequencing

Single-cell RNA-sequencing (scRNAseq) was performed on peripheral blood mononuclear cells (PBMC) from a subset of close contacts. PBMC were thawed, washed twice with phosphate buffered saline (PBS), and resuspended at 10E6 cells/100 ml in RPMI (MT10040CV from Mediatech/Corning) with 10% human serum. Cells were rested 2 hours, CD40 blocking antibody was added for 15 minutes (1:200 dilution, 0.5 ul), and then CD40L (diluted 1:10) was added. Cells were incubated overnight, washed with PBS, and then washed with cell staining buffer. Cells were resuspended in 50 ml of staining buffer and 5ml of the Fc blocking agent Human true stain FcX (cat#422302, Biolegend) and placed on ice for 10 minutes. Cells were hash-tagged using one microliter of TotalSeq-C Human Universal Cocktail (Biolegend) per sample and incubated on ice for a further 30 minutes. Cells were then washed three times and resuspended in staining buffer. A 137 Cite-seq C antibody (cat#399905, Biolegend) panel was prepared as per the manufacturer’s guidelines. For pools of 4-5 samples, 50ml of the antibody panel was added, and cells were incubated on ice for 30 minutes. Cells were washed three times with PBS with 2% BSA and resuspended in PBS with 1% BSA. Approximately 8,000 cells per individual were provided to the VANTAGE sequencing core. Libraries were created following the 10X Genomics standard protocol (3-prime version 4), and sequencing was performed on an Illumina NovaSeq 6000 at VANTAGE.

Raw sequences (FASTQ) were processed using 10x Genomics Cell Ranger Multi v9.0.0 using 10x Genomics Cloud Analysis (66). Poor-quality cells, doublets, and high mitochondrial cells (> 10%) were removed. Genes not expressed in at least 150 cells were removed. Imputation was applied to the concatenated count matrix using Adaptively-thresholded Low Rank Approximation (ALRA) (67) in R v4.3.3 (49). Further data cleaning and annotation was performed using python v3.10.16. A consensus set of highly variable genes (HVG, n = 5000) were calculated on the imputed counts across project batches. Principal component analysis (PCA, n = 30) was done. The PCA result was submitted to the Harmony algorithm for integration (theta = 2) (68). K-nearest neighbors (kNN, n = 30) were found using the Harmony components. Leiden clustering (resolution = 3) was used to partition the kNN graph. Cell clusters were split into three groups: T and natural killer (NK) cells, myeloid cells, and B cells. Each subset was then analyzed separately by HVG, PCA, Harmony, kNN, and Leiden clustering. Uniform Manifold Approximation and Projection (UMAP) was used for visualization. Leiden clusters were annotated according to canonical marker expression profiles (Supplemental info 1). Subset-specific HVG n-genes were set as follows: T/NK (n = 4000), myeloid (n = 2500), and B (n = 2000). Harmony theta was set as follows: T/NK (n = 4), myeloid (n = 4), and B (n = 2). Leiden resolution was set as follows: T/NK = 2.5, myeloid = 1, and B = 1.

### Single-cell expression quantitative loci (sceQTL)

In total, 57 participants with both scRNAseq and GWAS data were selected. Of these, 32 were close-control controls and 25 were enrolled as controls but progressed to PTB during the course of the study. Progressors were not included in the original GWAS; thus, ancestry PCs and relatedness were re-calculated for this combined subset as described above. Relatedness values were highly correlated across controls processed in this subset versus the full cohort (Pearson R = 0.998, P < 2.2E-16). sceQTL analysis was performed in R v4.4.2 (49). Single-cell data (543,229 cells) were pseudo-bulked using Seurat v5.1.0 (69), and the 32 cell types were processed independently. Counts were filtered to protein-coding genes, those with expression in at least 3 samples, trimmed mean of means (TMM) normalized, and transformed to log2 counts per million (CPM) with gene-level quality weights using edgeR (70) and limma (71, 72). This resulted in 7,297 to 13,029 genes for analysis per cell type. SNP data were filtered to overlapping single-cell participants and GWAS-significant SNPs that were annotated to at least one protein-coding gene, concordant with high-pass data, and expressed in at least one cell type in single-cell data. This yielded 8 SNPs annotated to 13 protein-coding genes, which resulted in 13 unique gene-SNP pairs. These pairs were modeled against gene expression in the 32 single-cell RNAseq cell types using a kimma (73) additive model. Covariate inclusion was assessed in univariate and leave-one-out models as with GWAS model fitting (Supplemental Table 6), which resulted in a final sceQTL additive model without clinical or PTB risk covariates (Supplemental Figure 7).

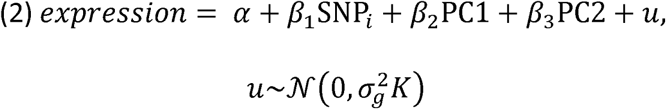

where components are as described in equation (1), except expression is the pseudo-bulk gene expression per individual within cell type. FDR correction was applied within cell type across SNPs annotated to the same gene. Significance was determined at FDR < 0.05. Significant sceQTLs were compared to the Database of Immune Cell eQTLs (DICE) (25) including matched or similar cell types, SNPs within 10 kb of significant sceQTLs reported here, and DICE P < 0.05.

Sensitivity analyses were performed including covariates relevant to GWAS models (sex, age, smoking, DM as in equation (1)) as well as for the interaction of SNP and PTB progression. The interaction term was not significant for any sceQTL (FDR > 0.05); thus, PTB progression was not considered in the final model. Dominant models were also assessed for SNPs with a measured but rare homozygous uncommon genotype (RGS9); all other SNPs lacked homozygous uncommon genotypes in the sceQTL subset. Finally, co-localization of GWAS significant SNPs and sceQTL signals were assessed by modeling all SNPs within 10 kb of the GWAS significant SNP against expression of the gene annotated to the GWAS SNP. SNPs at MAF > 1% were included similar to GWAS locus-zoom plots.

### curatedTBData bulk RNAseq analysis

Utilizing the curatedTBData R package (24), protein-coding genes associated with significant GWAS SNPs were assessed for association to PTB in bulk RNAseq or microarray. Of the 50 curated studies, 9 whole blood studies were selected based on the following criteria: TB negative controls, all HIV negative participants, at least 10 PTB participants, and expression of at least one gene of interest. For each study, normalized expression was subset to baseline samples from PTB and control participants only. Gene expression was modeled utilizing a simple linear model in kimma with covariates age and sex when available. Significance was defined at FDR < 0.05.

### Statistics

In the GWAS, an additive model was used and genotypes with < 3 homozygous uncommon individuals were confirmed by dominant model. GWAS P-values less than 5E-8 were considered significant and less than 5E-5 were suggestive. In sceQTL analysis, adjusted P-values less than 0.05 were considered significant and the Benjamini-Hochberg FDR was applied within cell type across SNPs annotated to the same gene. In gene enrichment, FDR less than 0.3 with a gene set overlap > 1 were considered significant. All summary values are mean ± standard deviation unless otherwise noted.

### Study approval

This study was approved by the institutional review boards of the study sites in Rio de Janiero (Instituto Nacional de Infectologia Evandro Chagas, Fundação Oswaldo Cruz [FIOCRUZ] [7.257.671], Faculdade de Medicina e Centro Municipal de Saúde de Duque de Caxias [7.294.692], Universidade Federal do Rio De Janeiro, Secretaria Municipal de Saúde do Rio De Janeiro e Clínica Da Família Rinaldo Delamare, [3.650.150]), Salvador (Instituto Brasileiro Para Investigação da Tuberculose [IBIT], Fundação José Silveira, [7.276.533]), and Manaus (Fundação de Medicina Tropical Dr. Heitor Vieira Dourado, [7.269.991]) as well as Vanderbilt University Medical Center (141049). Written informed consent was obtained from all participants.

## Data availability

Sequence files for this project are available in the database of Genotypes and Phenotypes (dbGaP phs004035.v1). All scripts associated with this project can be found in https://github.com/hawn-lab/ReporTB_GWAS_public. Data associated with Figure 1-4 can be found in the Supporting data values. Data for Figure 1, C and D as well as Figure 2A are filtered to P < 0.1.

## AUTHOR CONTRIBUTIONS

KADM, BBA, TRS, and TRH conceived the study with contributions from SAM, GA, RDG, and SAK. Funding and reagents were provided by BBA, SAK, TRS, and TRH. MCF and AMSA contributed to operational, technical, laboratory support, or project management. MCS, ALK, and VCR were responsible for all site-level activities, including recruitment, clinical management, and data collection. Experiments were conducted by FAR, JMCA, and JDS, and data were verified by KADM, JDS, and MCF. KADM led data analysis with contributions from JDS, JMO, JPA, and HIN. KADM, BBA, TRS, TRH, SAM, GA, and PFR contributed to results interpretation. KADM wrote the primary manuscript. All authors provided feedback on the manuscript and approved the final manuscript for submission. Co-first author order was determined by relative contributions to manuscript writing. Co-corresponding authors were designated given the complexity of the study design and implementation.

## ACKNOWLEDGEMENTS

We thank the individual study participants of the Regional Prospective Observational Research in Tuberculosis (RePORT)-Brazil consortium, study coordinators, and the clinical and research staff at all sites. See Supplemental Acknowledgments for details on RePORT-Brazil consortium. We would also like to thank Alexander Bick (VUMC) for his assistance and insights.

## SUPPLEMENTAL ACKNOWLEDGMENTS

### The Regional Prospective Observational Research in Tuberculosis (RePORT)-Brazil Consortium

<u>Salvador, BA, Brazil</u>

Laboratório de Pesquisa Clínica e Translacional, Instituto Gonçalo Moniz, Fundação Oswaldo Cruz (FIOCRUZ): Mariana Araújo-Pereira

Instituto de Pesquisa Clínica e Translacional, Medicina Zarns, Clariens Educação: Mariana Araújo-Pereira, Artur T L Queiroz

Instituto Brasileiro para Investigação da Tuberculose, Fundação José Silveira: Michael Rocha, Vanessa Nascimento, Victor Porfirio

<u>Rio de Janeiro, RJ, Brazil</u>

Programa Acadêmico de Tuberculose da Faculdade de Medicina, Universidade Federal do Rio de Janeiro: Adriana Moreira, André Luiz Bezerra, Anna Cristina Carvalho

Laboratório de Pesquisa Clínica em Micobacteriose, Instituto Nacional de Infectologia Evandro Chagas (FIOCRUZ): Adriano Gomes Silva, Aline Benjamin, Quezia Medeiros, Flavia M Sant’Anna

Secretaria Municipal de Saúde do Rio De Janeiro e Clínica Da Família Rinaldo Delamare: Solange Cavalcante, Betina Durovni, Jamile Garcia, João Marine

<u>Manaus, AM, Brazil</u>

Fundação de Medicina Tropical Dr. Heitor Vieira Dourado do Amazonas: Allyson G Costa, Brenda Carvalho, Alexandra Brito, Leandro Garcia, Renata Spener

<u>Nashville, TN, USA</u>

Division of Infectious Diseases, Department of Medicine, Vanderbilt University Medical Center: Angela Jones, Hilary V Riley, Cody Staats, Megan Turner

<u>Seattle, WA, USA</u>

Division of Allergy and Infectious Diseases, Department of Medicine, University of Washington: Josh J Ivie

