## Supplementary Figures and Table legends for "Genome-wide association study in Brazil identifies genetic susceptibility to tuberculosis with single-cell gene effects"

**SUPPLEMENTARY MATERIAL**

**Supplemental Table 1. Concordance and quality of imputed low-pass and 30X high-pass SNPs**. Total participants out of 50 with a pass-filter call in 30X high-pass data, the percent of these calls that match between low- and high-pass data, imputation scores, and minor allele frequency (MAF) are reported for pass-filter SNPs with concordance > 75%, imputation score > 0.6, and MAF > 1%.

**Supplemental Table 2. GWAS model results**. Unless otherwise noted, all model results include model terms, estimates, p-values, Akaike information criterion (AIC) fit, and odds-ratios (OR) for all pass-filter SNPs at 5% minor allele frequency (MAF) and 75% concordance with high-pass sequencing. The full cohort full model was TB ~ SNP + PC1 + PC2 + sex + age + smoking + HIV + DM + cov(coefficient of relatedness). Models for the risk-controlled sub-cohort excluded HIV and DM as all sub-cohort participants were HIV and DM negative. Models with no TB risks excluded smoking, HIV, and DM. Univariate and leave-one-out model fitting were completed using the full cohort with a random subset of 10,000 SNPs and only AIC fit is reported. Models utilizing the 3-level smoking covariate (never, past, current) present only the P-values for current vs past and current vs never for the 10,000 SNP subset. As current and past showed no significant differences (P > 0.31), the 2-level smoking covariate was used in the final model (current or past vs never). Covariate outcome models replaced the TB outcome with one clinical or TB risk covariate. These were run separately in case and control subsets for SNPs significant (P < 5E-8) in the full cohort. MAF 1% models were run in the full cohort and include SNPs within 10 kb of significant GWAS SNPs. These models report score and full model P-values as well as linkage disequilibrium (LD) R^2^ to the lead significant GWAS SNP.

**Supplemental Table 3. TB disease heritability estimates with and without covariates.** GCTA was used to estimate TB disease heritability with and without correction for ancestry principal components (PC), age, sex, and TB risk factors including smoking, HIV, and DM. VG: genetic, Ve: environmental, Vp: phenotypic (VG+Ve), Vp_L: scaled phenotypic

**Supplemental Table 4. Significant GWAS results and annotations**. SNPs significant (P < 5E-8) in the full cohort are included (N = 17). The percent concordance is given for each SNP called in imputed low-pass data versus 30X high-pass data (hpWGS_concordance) and low-pass data versus Twist targeted sequencing (Twist_concordance). P-values and odds-ratios (OR) are reported for both the full and risk-controlled (cc) cohorts. Minor allele frequency (MAF) and total counts (N) of homozygous common (0), heterozygous (1), and homozygous uncommon (2) are given in cases and controls. Annotations are given for intragenic and 50 kb cis genes. HUGO symbols are reported and if no HUGO symbol exists for a gene, the ENSEMBL ID is given. Genes have up to 2 intragenic and 10 cis annotations. An additional sheet provides the P-values for full cohort models excluding one or more covariates.

**Supplemental Table 5. Significant pathway enrichment of protein-coding genes associated with GWAS significant and concordant SNPs**. Genes were enriched against Broad MSigDB Hallmark, canonical pathways, and gene ontology. Gene sets with FDR < 0.3 are reported. =k_median: overlap of significant genes in gene set, median calculated across 100 random subsets to one annotation per SNP; K: total size of gene set.

**Supplemental Table 6. Single-cell eQTL model results.** Unless otherwise noted, all model results include model terms, estimates, p-values, and Akaike information criterion (AIC) fit for SNPs significant in the full cohort GWAS (P < 5E-8) and annotated to a protein-coding gene. The final model was expression ~ SNP + PC1 + PC2 + cov(coefficient of relatedness). Models with additional covariates were expression ~ SNP + PC1 + PC2 + sex + age + smoking + DM + cov(coefficient of relatedness). Univariate and leave-one-out model fitting were completed, and AIC fit is reported. Cell cluster short and long annotations are provided.

**Supplemental Table 7. Overlap significant GWAS SNPs with SNP arrays used in prior TB GWAS studies**. The 17 full cohort significant, concordant SNPs were compared to SNPs present on 19 arrays from Affymetrix or Illumina. Previous GWAS study methods and significant results are summarized with the GWAS results of this study reported for prior significant SNPs. In addition, the 17 SNPs found in this study were searched in past TB GWAS with all but one not reporting results for comparison due to quality, significance, or other filters applied to reported P-value tables.

**Supplemental Table 8. Differential gene expression for select curatedTB bulk RNAseq datasets**. Protein-coding genes associated with significant GWAS SNPs were assessed for association to PTB in nine whole blood bulk RNAseq or microarray datasets available in curatedTB. Linear model results are reported per study as well as summarized at FDR < 0.05.

**Supplemental Table 9. DICE result comparison**. Significant sceQTLs were compared to the Database of Immune Cell eQTLs (DICE) including matched or similar cell types, SNPs within 10 kb of significant sceQTLs reported here, and DICE P < 0.05.

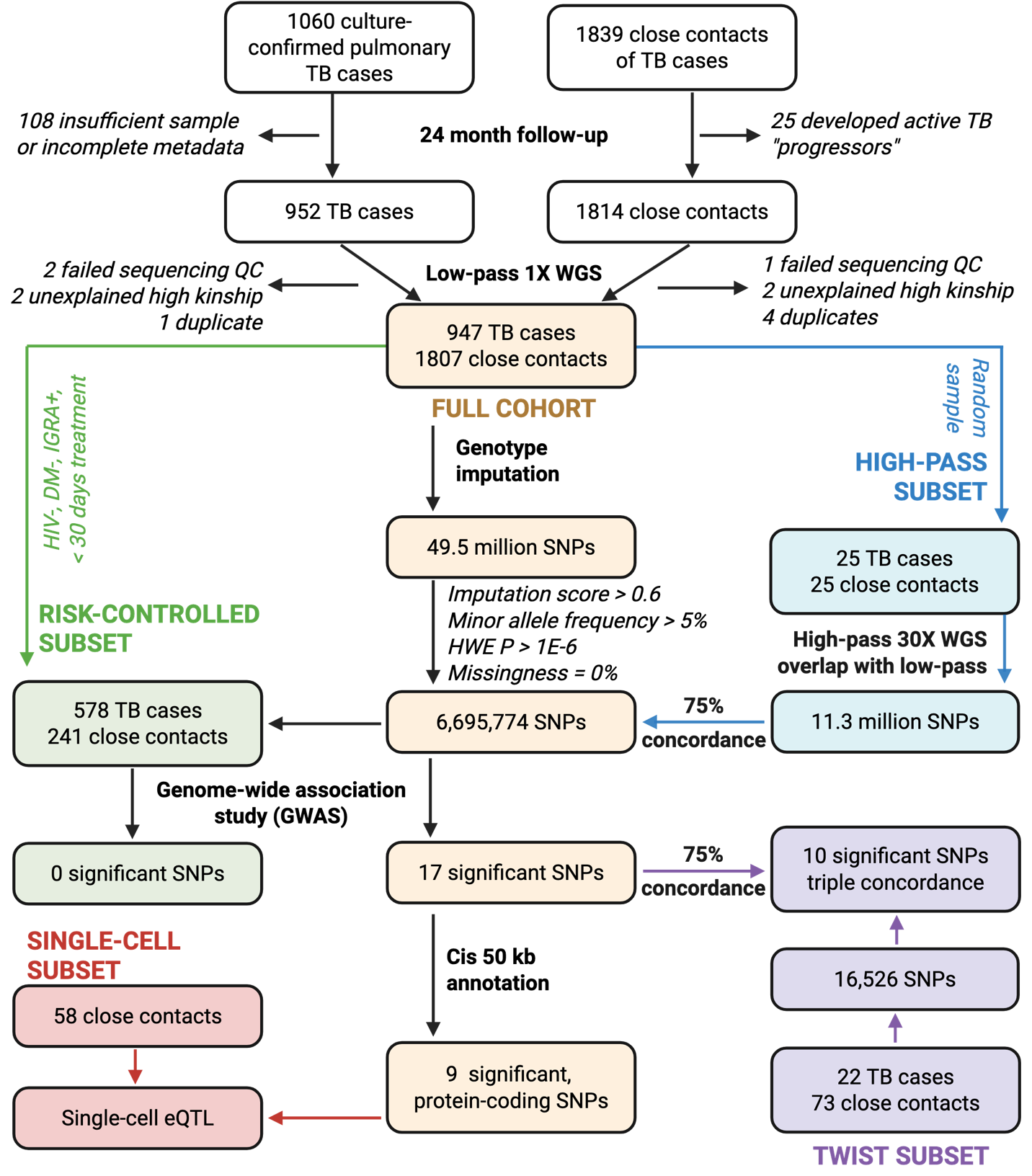

**Supplemental Figure 1. Detailed GWAS study and analysis design.** Tuberculosis (TB) cases and their close contacts were recruited as part of RePORT-Brazil. GWAS was performed using 1X low pass whole genome sequencing (lpWGS) with imputation. The full cohort (N = 2754) and a risk-controlled sub-cohort controlling for HIV, diabetes, and LTBI treatment (N = 819) were assessed separately. High-pass 30X WGS was completed for a subset of 50 participants, and SNPs were filtered for low-pass quality (imputation score, minor allele frequency [MAF], Hardy-Weinberg equilibrium [HWE], missingness, and > 75% concordance with high-pass genotype calls. In total, 17 SNPs were significant in the full cohort with 8 annotated to at least one protein-coding gene up to 10 kb cis. . These 8 significant protein-coding SNPs were assessed in single cell expression quantitative trait loci (sceQTL) analysis of 58 participants. Finally, 10 significant, concordant SNPs were further verified by > 75% concordance with targeted probe-based Twist sequencing of 95 participants.

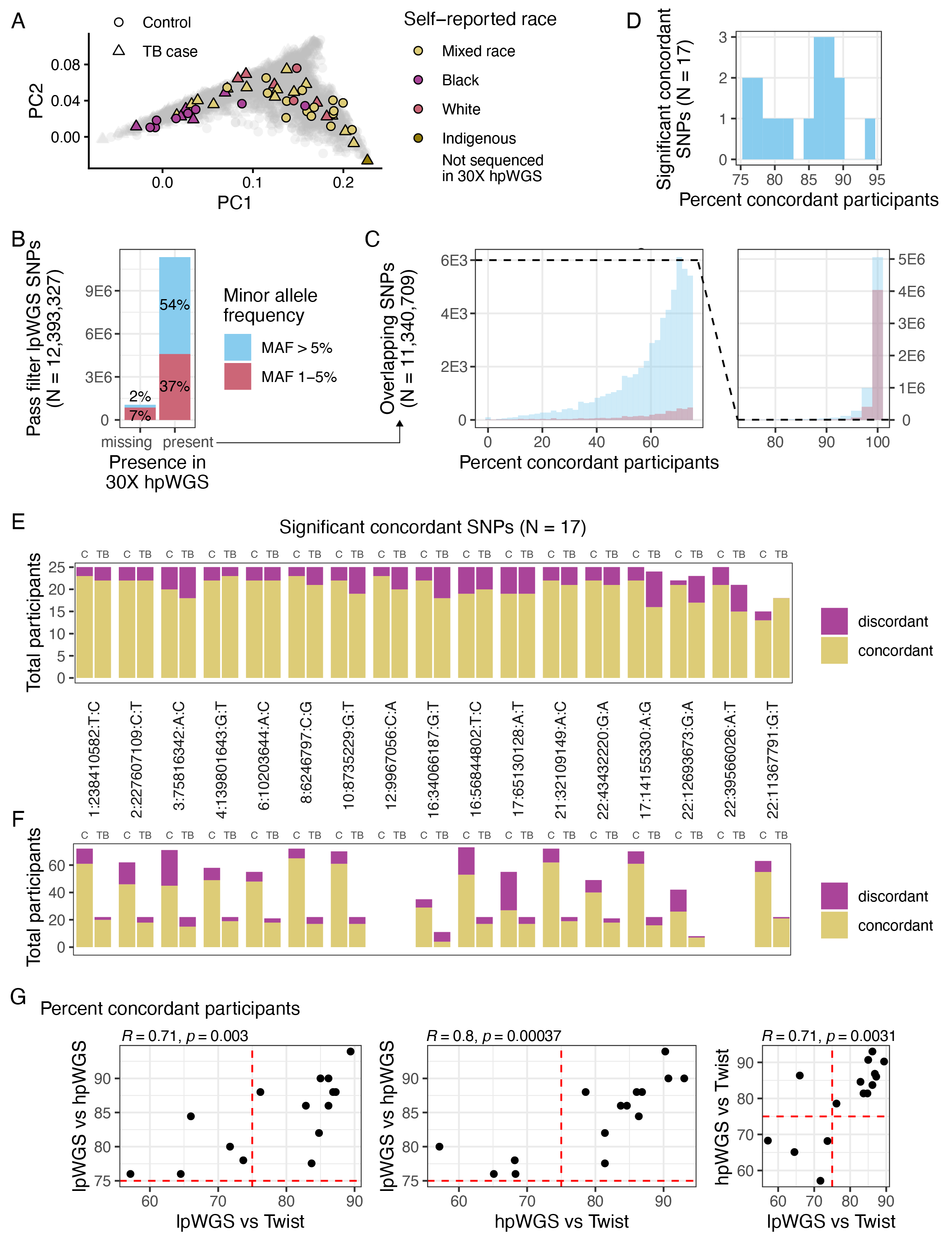

**Supplemental Figure 2**. **High-pass 30X whole genome sequencing confirmation of imputed low-pass genotypes**. A subset of 50 participants was sequenced with 30X high-pass WGS (hpWGS). (**A**) Principal component analysis (PCA) with SNPs also found in 1000 Genomes highlighting individuals included in hpWGS (N = 139,855 SNPs). RePORT-Brazil individuals are stratified by self-identified ancestry most closely matched to 1000 Genomes Super Populations as in **Supplemental Figure 2**. (**B**) Call rate of imputed, pass-filter (score > 0.6, MAF > 1%, HWE P > 1E-6, 0% missingness) low-pass WGS (lpWGS) SNPs (N = 12.4 million) in hpWGS data. Color indicates minor allele frequency (MAF) of 1% (locus zoom analysis) and 5% (GWAS). (**C**) Percent concordance of overlapping SNPs present in both lpWGS and hpWGS (N = 11.3 million SNPs). Concordance is split at the applied cutoff as the majority of pass-filter lpWGS SNPs were above 75% concordance (99.5%). The dashed line indicates equivalent y-axes values. In this visualization, SNPs were not filtered for minimum total overlapping calls. (**D**) Percent concordance of GWAS significant SNPs filtered to > 75% and a minimum of 20 overlapping genotype calls between lpWGS and hpWGS (N = 17). Concordance between (**E**) lpWGS and hpWGS (control [C], TB cases [TB], N = 25 each) or (**F**) lpWGS and Twist probe-based sequencing (C N = 73, TB N = 22) for significant, concordant SNPs (N = 17 SNPs). Bars are colored by the total participants with calls concordant or discordant between two data sets. Two SNPs were not called in Twist data. (**G**) Pearson correlation of lpWGS, hpWGS, and Twist concordance.

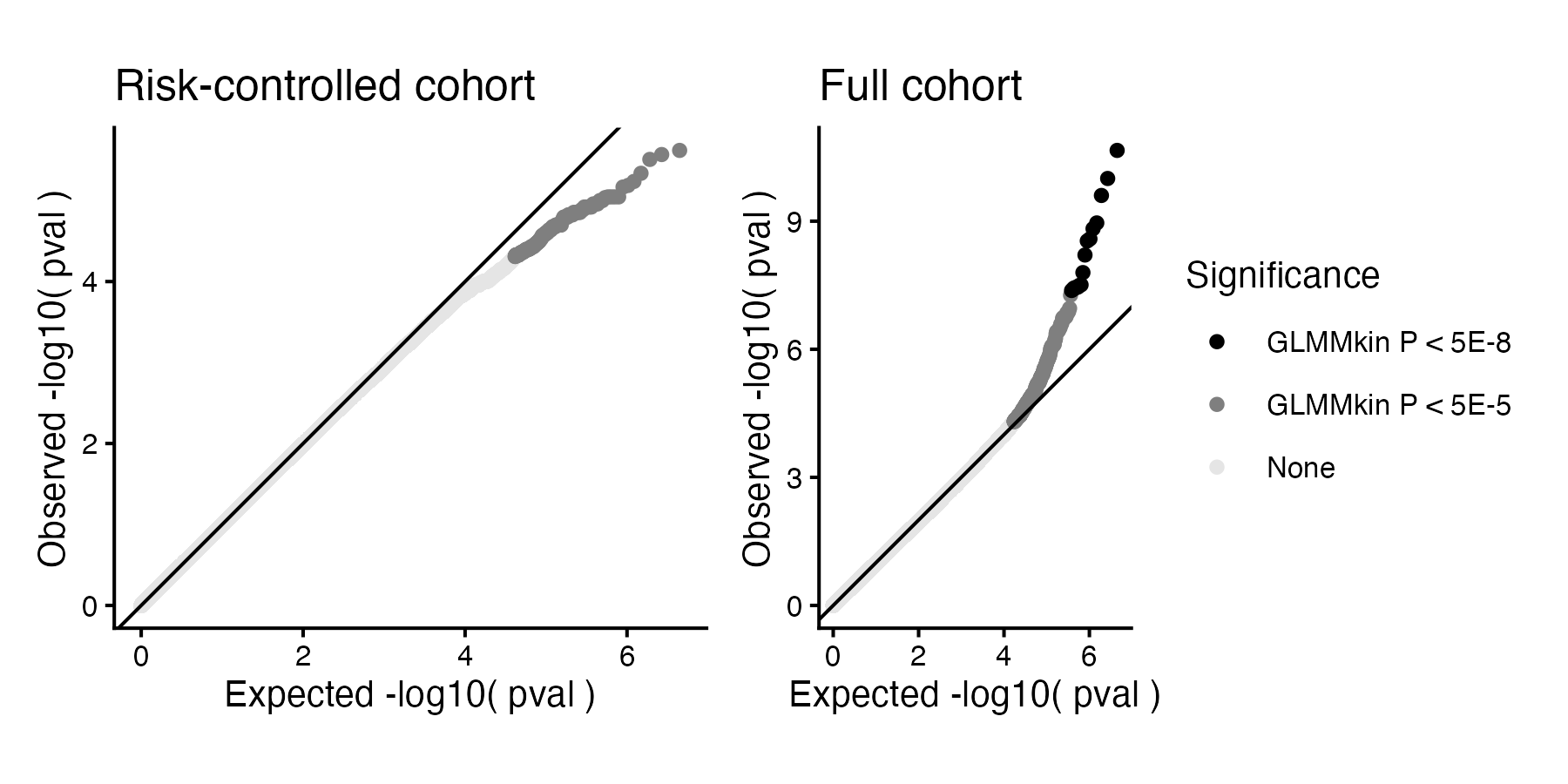

**Supplemental Figure 3. Genome-wide QQ plot in the full cohort and risk-controlled sub-cohort**. Shading indicates genotypes below the GLMMkin full model suggestive P < 5E-5 or significant P < 5E-8. The solid line indicates a 1:1 ratio between the expected and actual p-values. Genomic inflation factors (λ) are 0.976 for the risk-controlled and 0.998 for the full cohort.

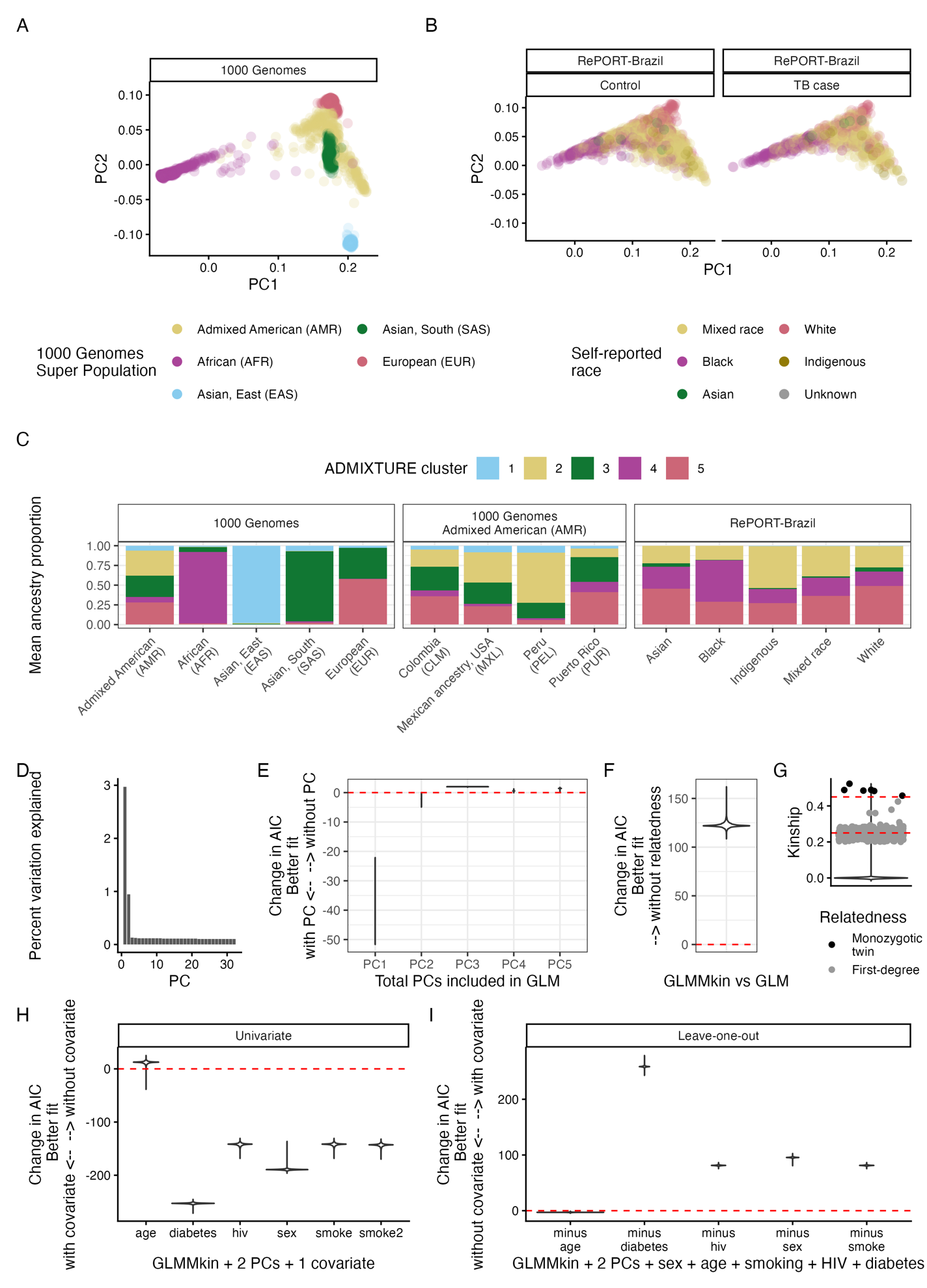

**Supplemental Figure 4**. **GWAS model assessment with ancestry PCs, genetic relatedness, and clinical covariates.** Genotypes present in both datasets were combined and filtered for RePORT-Brazil and 1000 Genomes phase 3 (N = 139,855 SNPs). Principal component analysis (PCA) revealed overlap of several (**A**) 1000 Genomes Super Populations and (**B**) RePORT-Brazil individuals. RePORT-Brazil individuals were stratified by self-identified ancestry most closely matched to 1000 Genomes Super Populations in PCA space. (**C**) Predicted ancestry proportions (K = 5) in 1000 Genomes Super Populations, 1000 Genomes Admixed American populations, and RePORT-Brazil. ADMIXTURE clusters are colored by the Super Population in which they have the highest proportion. (**D**) Percent variation of the entire genetic dataset explained by each ancestry PC. Randomly selected genotypes were then modeled for TB cases vs controls (N = 10,000 SNPs). Model fit with (**E**) the addition of ancestry PCs or (F) the coefficient of relatedness (2 x kinship). For (E), change in AIC was calculated as the model with N+1 PCs minus the model with N PCs where for (**F**), change in AIC was a model with 2 PCs +/- relatedness. Negative AIC indicates improved model fit with the additional covariate. Plots are violin summaries across 10,000 genotypes. (**G**) Pairwise genetic kinship. Each dot represents a pair of samples. All pairs are summarized by violin plot with red dashed lines indicating mean expected kinship for monozygotic twins. Additionally, pairs with kinship > 0.2 (first-degree) are shown in dots with color indicating monozygotic twins (black) and potential first-degree relatives (grey). (**H**) Model fit with the addition of a single clinical covariate including age, diabetes, HIV, sex, and smoking (three-level and two-level). Negative change in AIC indicates improved fit with the additional covariate. (**I**) Model fit with the removal of a single clinical covariate from the full model. Positive change in AIC indicates worse model fit with the removal of the covariate.

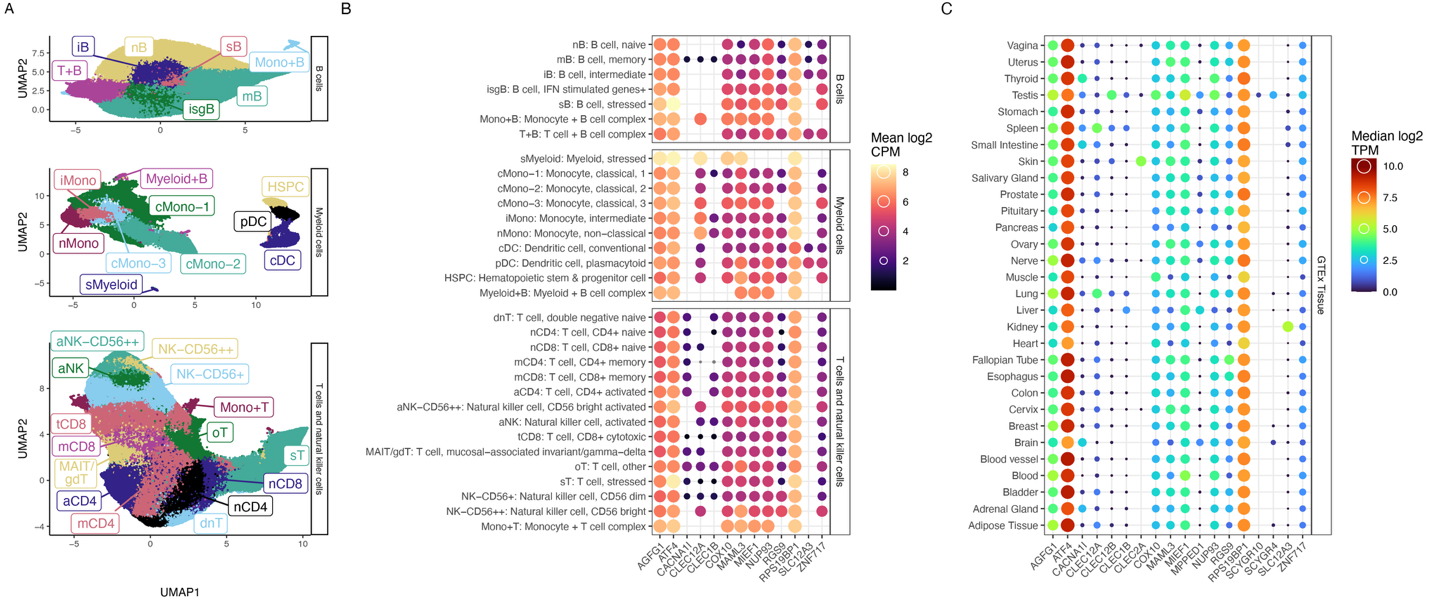

**Supplemental Figure 5.** **UMAP dimension reduction and gene expression of genes associated with GWAS significant SNPs**. GWAS significant concordant SNPs were mapped 50 kb cis to protein-coding genes. Gene expression was modeled for corresponding mapped SNPs. (**A**) UMAPs depict cells labeled by cell type (N = 32) split by B cells, myeloid cells, and T and natural killer cells. Labels are described in B with cell type assignment details in **Supplemental info 1**. (**B**) Expression is shown for GWAS associated genes expressed in at least one of the 32 cell types (N = 8 SNPs, 13 genes). All cell types identified in single-cell RNAseq are shown in roughly the same order as (A). . Color and size indicate mean log2 counts per million (CPM) of each gene in each cell type. Blanks represent genes that did not pass abundance filters for analysis in that cell type (CPM > 0 in at least 3 samples). (**C**) Expression of GWAS associated genes expressed in at least one tissue in the Genotype-Tissue Expression (GTEx) Portal (N = 18) regardless of single-cell RNAseq expression in (B). Two genes were not expressed in any GTEx tissue (SCYGR2,3) and are not depicted here. Color and size indicate median log2 transcripts per million (TPM).

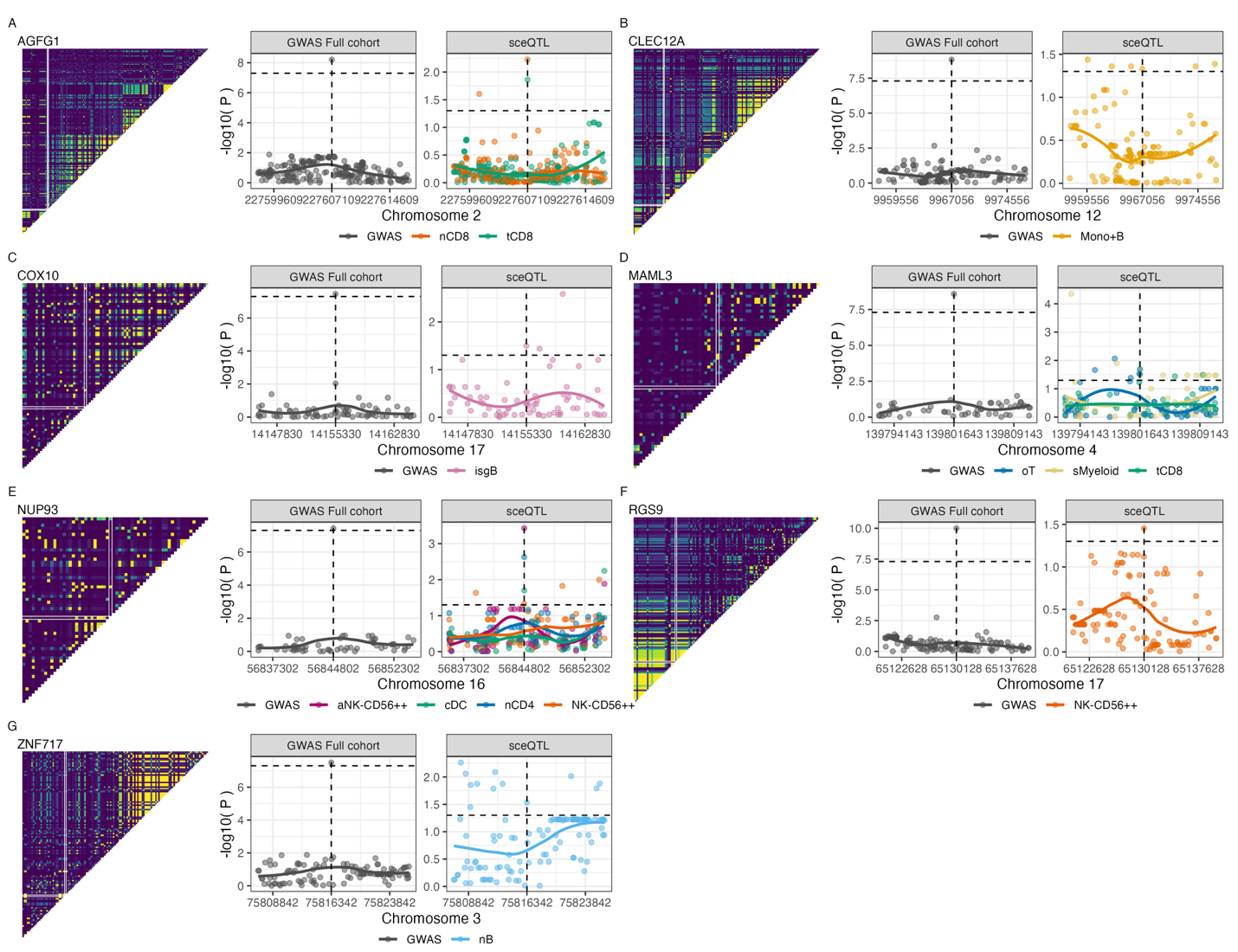

**Supplemental Figure 6. Linkage disequilibrium and co-localization of sceQTLs.** SNPs at MAF > 1% and +/- 10 kb from the lead SNP were selected for (**A**) AGFG1 (2:227607109:C:T, downstream), (**B**) CLEC12A (12:9967056:C:A, intron), (**C**) COX10 (17:14155330:A:G, intron), (**D**) MAML3 (4:139801643:G:T, intron), (**E**) NUP93 (16:56844802:T:C, intron), (**F**) RGS9 (17:65130128:A:T, intron), and (**G**) ZNF717 (3:75816342:A:C, downstream). (Left) Linkage disequilibrium (R^2^) was calculated for all pairwise comparisons. The significant SNP position is outlined in white. (Right) Co-localization of GWAS and sceQTL signals. Significance trends are plotted using LOESS fits colored by cell type. Horizontal dashed lines indicate significance (P < 5E-8 GWAS, P < 0.05 sceQTL). See Figure S6 for cell type annotations.

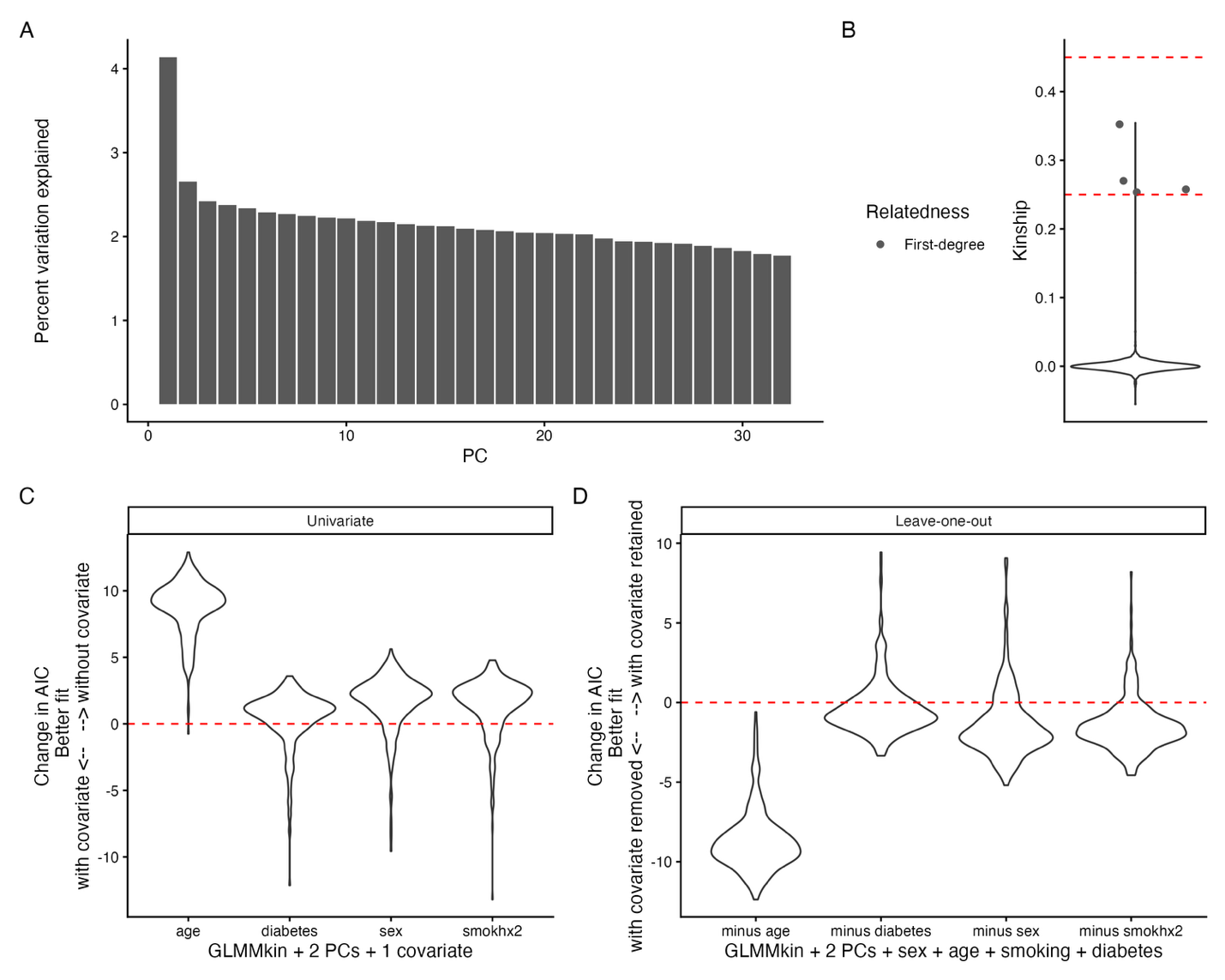

**Supplemental Figure 7.** **Single-cell eQTL model assessment with ancestry PCs, genetic relatedness, and clinical covariates.** In total, 58 participants were assessed for 8 SNPs significantly associated with TB (P < 5e-8) and annotated to protein-coding genes expressed in at least one cell type in single-cell RNAseq. SNPs were modeled against expression of their corresponding gene(s) in individual cell types. (**A**) Percent variation of the entire genetic dataset explained by each ancestry PC. (**B**) Pairwise genetic kinship. Each dot represents a pair of samples. All pairs are summarized by violin plot with red dashed lines indicating mean expected kinship for twins and first-degree relatives. Additionally, pairs with kinship > 0.2 (third-degree) are shown in dots with color indicating potential first-degree relatives (grey). No monozygotic twins were present in this dataset. . (**C**) Model fit with the addition of a single clinical covariate including age, diabetes, sex, and smoking (two-level). Negative change in AIC indicates improved fit with the additional covariate. (**D**) Model fit with the removal of a single clinical covariate from the full model. Positive change in AIC indicates worse model fit with the removal of the covariate.

**Supplemental Info 1. Single-cell annotations markers.**

T and NK cells

CD4+ T Memory:

coarse RNA: CD3G+CD74-CD14-NKG7-CD19-LYZ- [qualifies as T cell]

coarse ADT: CD3+CD56-CD14-CD1c-CD19- [qualifies as T cell]

subset RNA: CD3E+CD4+CD8A-CCR7- [qualifies as CD4 T Memory]

subset ADT: CD45RO+CD45RA- [further evidence of a Memory phenotype]

CD4+ T Naive:

coarse RNA: CD3G+CD74-CD14-NKG7-CD19-LYZ- [qualifies as T cell]

coarse ADT: CD3+CD56-CD14-CD1c-CD19- [qualifies as T cell]

subset RNA: CD3E+CD4+CD8-CCR7+ [qualifies as CD4 T Naive]

subset ADT: CD45RO-CD45RA+ [further evidence of a Naive phenotype]

CD8+ T Cytotoxic:

coarse RNA: CD3G+CD74-CD14-CD19-LYZ- [qualifies as T cell]

coarse ADT: CD3+CD56-CD14-CD1c-CD19- [qualifies as T cell]

subset RNA: CD3E+CD4-CD8A+PRF1+GNLY+CCR7- [perforin, granulysin with CD8 T memory phenotype is consistent with a cytotoxic label]

subset ADT: GPR56+CD57+KLRG1+ suggests a more terminally differentiated effector memory type consistent with a CD8+ T cytotoxic label

CD8+ T Memory:

coarse RNA: CD3G+CD74-CD14-CD19-LYZ- [qualifies as T cell]

coarse ADT: CD3+CD56-CD14-CD1c-CD19- [qualifies as T cell]

subset RNA: CD3E+CD4-CD8A+CCR7-LEF1- [qualifies as a CD8 T Memory]

subset ADT: overall pattern is consistent with transcript-level annotation

CD8+ T Naive:

coarse RNA: CD3G+CD74-CD14-CD19-LYZ- [qualifies as T cell]

coarse ADT: CD3+CD56-CD14-CD1c-CD19- [qualifies as T cell]

subset RNA: CD3E+CD4-CD8A+CCR7+LINC02446+ [qualifies as CD8 T Naive]

subset ADT: CD45RO-CD45RA+ [further evidence of a Naive phenotype]

dnT Naive: dnT = CD4-CD8- T cell

coarse RNA: CD3G+CD74-CD14-CD19-LYZ- [qualifies as T cell]

coarse ADT: CD3+CD56-CD14-CD1c-CD19- [qualifies as T cell]

subset RNA: CD3E+CD4lowCD8lowCCR7+LEF1+LINC02446+ [qualifies as a Naive type without clear CD4 or CD8 assignment]

subset ADT: CD3+CD8lowCD4lowCD45RO-CD45RA+ [further evidence that dnT Naive is appropriate]

MAIT/gdT:

coarse RNA: CD3G+CD74-CD14-CD19-LYZ- [qualifies as T cell]

coarse ADT: CD3+CD56-CD14-CD1c-CD19- [qualifies as T cell]

subset RNA: CD3E+CD4-KLRB1+TRGV9+TRAV1-2 [qualifies as a mixed MAIT, gdT group]

subset ADT: TCR.Vd2+TCR.AB-(characteristic of gdT)CD161+(MAIT marker)CD45RO+CD45RA-(non-naive) [all consistent with mixed MAIT, gdT label]

Other T:

coarse RNA: CD3G+CD74-CD14-CD19-LYZ- [qualifies as T cell]

coarse ADT: CD3+CD56-CD14-CD1c-CD19- [qualifies as T cell]

subset RNA: CD3E- relative to other T cells but CD4+ suggests a non-classical type

subset ADT: unclear

Stress T:

https://doi.org/10.1016/j.isci.2023.107588 [describe a similar HSP cluster]

coarse RNA: CD3G+CD74-CD14-CD19-LYZ- [qualifies as T cell]

coarse ADT: CD3+CD56-CD14-CD1c-CD19- [qualifies as T cell]

subset RNA: T cell from coarse markers RNA; CD69+HSPA6+HSPA2+ consistent with the Stress label (equivalent to other paper's HSP T)

subset ADT: CD4 T cell from coarse markers ADT; subset heatmap does not add value

Tcell:Mono:

coarse RNA: CD3,CD14 mixed signal (CD3GlowCD14+) [partitioned with T cells]

coarse ADT: CD3,CD14 mixed signal (CD3lowCD14low) [partitioned with T cells]

subset RNA: does not add value but is consistent

subset ADT: does not add value but is consistent

Activated NK:

coarse RNA: NKG7+CD3G-CD4-CD19-CD74-CD14- [qualifies as NK]

coarse ADT: CD56+CD3-CD19-CD1c-CLEC12A- [consistent with NK label]

subset RNA: CD38+FCGR3A+KLRB1+NCAM1dimXCL1- [consistent with a CD56dim activated type; CD38+NCAM1dim is the key signature]

subset ADT: CD38+CD16+CD161+CD56dim (relative to other NK) is consistent with gene signature. FCGR3A=CD16 gene; KLRB1=CD161 gene; NCAM1=CD56 gene

NK CD56bright:

coarse RNA: NKG7+CD3G-CD4-CD19-CD74-CD14- [qualifies as NK]

coarse ADT: CD56+CD3-CD19-CD1c-CLEC12A- [consistent with NK label]

subset RNA: XCL1+NCAM1bright [NK CD56bright hallmark]

subset ADT: CD56brightCD94+CD314+ with low CD158(KIR) family profile [supports the NK CD56bright label]

NK CD56dim:

coarse RNA: NKG7+CD3G-CD4-CD19-CD74-CD14- [qualifies as NK]

coarse ADT: CD56+CD3-CD19-CD1c-CLEC12A- [consistent with NK label]

subset RNA: XCL1-NCAM1dim [NK CD56dim hallmark]

subset ADT: CD56dimCD314-D94- with high/diverse CD158(KIR) family profile [supports the NK CD56dim label]

Myeloid cells

cDC:

coarse RNA: NKG7-CD3G-CD8A-CD19-CD74+CD14-S100A8- [supports non-Monocyte myeloid type]

coarse ADT: CD3-CD56-CD19-CLEC12A+ [supports myeloid type]

subset RNA: AFF3+HLA_DQA1+ENHO+ [markers of cDC, specifically cDC2]

subset ADT: CD14-CD16-CD1c+FCER1A+CD11b- [consistent with DC]

cMono:

coarse RNA: NKG7-CD3G-CD8A-CD19-CD74-CD14+S100A8+ [supports Monocyte myeloid type]

coarse ADT: CD3-CD56-CD19-CLEC12A+ [supports myeloid type]

subset RNA: CD14+S100A8+FCGR3Adim [supports a classical Monocyte label]

subset ADT: CD14+CD16dim [supports a classical Monocyte label]

HSPC:

coarse RNA: NKG7-CD3G-CD8A-CD19-CD74+CD14-S100A8- [supports non-Monocyte myeloid type]

coarse ADT: CD3-CD56-CD19-CLEC12A+ [supports myeloid type]

subset RNA: CD14-S100A8-FCGR3A-CD34+MYB+LAPTM4B+KIT+ [defines HSPC]

subset ADT: does not add value

pDC:

coarse RNA: CD3G-CD8A-CD19-CD74+CD14-S100A8- [supports non-Monocyte myeloid type]

coarse ADT: CD3-CD56-CD19-CLEC12A+ [supports myeloid type]

subset RNA: IRF4+MZB1+SMPD3+PLD4+TPM2+SPIB+CD14-CD34- [defines pDC]

subset ADT: CD123+CD14- [consistent with pDC]

iMono:

coarse RNA: NKG7-CD3G-CD8A-CD19-CD14+S100A8+ [supports Monocyte myeloid type]

coarse ADT: CD3-CD56-CD19-CLEC12A+CD14+S100A8+ [supports Monocyte myeloid type]

subset RNA: CD14+FCGR3A+ [consistent with intermediate Mono]

subset ADT: CD14+CD16+ with CD163+CD196+ supports an intermediate Mono label (transitional phenotype)

Stress Myeloid:

coarse RNA: NKG7-CD3G-CD8A-CD19-CD74-CD14+S100A8+ [supports Monocyte myeloid type]

coarse ADT: CD3-CD56-CD19-CLEC12A+ [supports myeloid type]

subset RNA: HSPA2+HSPA14+HSPA6+ with broad myeloid character supports the Stress Myeloid label

subset ADT: low CD14,CD16 dual protein expression with more CD14 transcript than the DC types suggests a potential non-canonical Monocyte or other character; the non-specific Stress Myeloid label is reasonable

Mono:B:

coarse RNA: NKG7-CD3G-CD8A-CD19+MS4A1+CD14+S100A8+ [defines Mono:B complex]

coarse ADT: CD3-CD56-CD19+CLEC12A+CD14+ [defines Mono:B complex]

subset RNA: CD19+MS4A1+CD14+S100A+ [consistent with Mono:B mixed signal]

subset ADT: does not add value (B cell markers seem nonspecific here, dropped from heatmap; use RNA signature for definition)

nMono:

coarse RNA: NKG7-CD3G-CD8A-CD19-CD14+ [supports Monocyte myeloid type]

coarse ADT: CD3-CD56-CD19-CLEC12A+ [supports myeloid type]

subset RNA: CD14dimFCGR3A+ [defines nonclassical Monocyte]

subset ADT: CD14dimCD16+ [defines nonclassical Monocyte]

B cells

B Intermediate:

coarse RNA: CD19+MS4A1+CD3G-CD4-CD8A-NKG7-CD14- [qualifies as B cell]

coarse ADT: CD19+CD20+CD3-CD4-CD56-CD14-CLEC12A- [qualifies as B cell]

subset RNA: intermediate for both memory and naive markers; see B Memory and B Naive [evidence that Intermediate label is reasonable]

subset ADT: consistent pattern. no real value added. CD27 dim consistent with non-memory label.

B Memory:

coarse RNA: CD19+MS4A1+CD3G-CD4-CD8A-NKG7-CD14- [qualifies as B cell]

coarse ADT: CD19+CD20+CD3-CD4-CD56-CD14-CLEC12A- [qualifies as B cell]

subset RNA: CD27+LINC01781+SSPN+AIM2+YBX3- [consistent with B Memory, particularly CD27]

subset ADT: CD27+ [consistent with memory]

B Naive:

coarse RNA: CD19+MS4A1+CD3G-CD4-CD8A-NKG7-CD14- [qualifies as B cell]

coarse ADT: CD19+CD20+CD3-CD4-CD56-CD14-CLEC12A- [qualifies as B cell]

subset RNA: CD27-LINC01781-SSPN-AIM2-YBX3+ [consistent with B Naive]

subset ADT: dim for CD27 [consistent with naive]

B:Tcell:

coarse RNA: CD19+MS4A1+CD14- [qualifies as B cell; shows some dim T cell character]

coarse ADT: CD19+CD20+CD3+CD4dimCD56-CD14-CLEC12A- [evidence for B:Tcell, likely CD4T, complex]

subset RNA: CD3E+ paired with coarse marker profile [evidence for B:Tcell]

subset ADT: CD3+ with low CD19 relative to true B cell types [consistent with B:Tcell]

ISG B:

coarse RNA: CD19+MS4A1+CD3G-CD4-CD8A-NKG7-CD14- [qualifies as B cell]

coarse ADT: CD19+CD20+CD3-CD4-CD56-CLEC12A- [qualifies as B cell]

subset RNA: ISG20+GBP(1,2,4,5)+STAT1+ defines the ISG tag; CD69+ also suggests activation

subset ADT: no added value

Stress B:

coarse RNA: CD19+MS4A1+CD3G-CD4-CD8A-NKG7-CD14- [qualifies as B cell]

coarse ADT: CD19+CD20+CD3-CD4-CD56-CD14-CLEC12A- [qualifies as B cell]

subset RNA: HSPA6+HSPA2+HSPA14+ with CD69 suggests a "stressed" state

subset ADT: no added value

B:Mono:

coarse RNA: CD19+MS4A1+CD14+S100A8+CD3G-NKG7- [evidence for B:Mono, likely cMono by FCGR3A, complex]

coarse ADT: CD19+CD14+CLEC12A+CD3-CD4-CD8-CD56- [evidence for B:Mono complex]

subset RNA: CD14+ [consistent with coarse marker profile and B:Mono label]

subset ADT: increased CD14 is consistent with B:Mono
